# Temperature and Low-stakes Cognitive Performance

**DOI:** 10.1101/2021.10.15.21265034

**Authors:** Xin Zhang, Xi Chen, Xiaobo Zhang

## Abstract

In this paper we offer among the first evidence in a developing country context that transitory exposure to heat waves may disrupt low-stakes cognitive activities across a wide range of age cohorts. Matching cognitive test scores from a nationally representative longitudinal survey in China with weather data according to the exact time and geographic location of assessment, we find that exposure to a temperature above 32 °C on the test date, relative to a day within 22–24 °C, leads to a sizable decline in math scores by 0.088 SDs (equivalent to 0.30 years of education). The negative effect is more pronounced for less educated older adults taking math tests. Test takers living in hotter regions or having air conditioning installed on site are less vulnerable to extreme high temperatures, indicating the role of adaptation.

**JEL Codes:** I24, Q54, Q51

## 1. Introduction

Climate change has brought more frequent extreme temperatures, such as heat waves and cold spells. The world’s average temperature has increased 0.6 °C in the past three decades and 0.8 °C in the past century, and the trend is projected to continue (Hansen et al. 2006). The Intergovernmental Panel on Climate Change (IPCC) warns that, if greenhouse gas emissions continue at the current rate, by 2034 the atmosphere may warm up by as much as 1.5 °C (2.7 °F) above the preindustrial levels (IPCC 2021). Along with rising temperature, heat waves are expected to occur more often.

There are several channels through which extreme temperatures may impede cognitive performance. First, cognitive activities often rely on regions in the brain sensitive to heat or cold weather, causing impaired brain functioning (Hocking et al. 2001; Kiyatkin 2007). Second, exposure to heat waves may reduce flow of blood to the brain (Kiyatkin 2007; Raichle and Mintun 2006) and therefore increase heat-related fatigue (McMorris et al. 2006; Nybo, Rasmussen, and Sawka 2014). Third, thermal stress may diminish respondents’ attention, working memory, information retention and processing (Hocking et al. 2001; Vasmatzidis, Schlegel, and Hancock 2002).

A growing body of literature assesses the impact of extreme temperatures, particularly heat waves, on cognitive performance. Some studies examine the effect of exposure to heat waves on students’ high-stakes exams (Cho 2017; Graff Zivin et al. 2020; Park 2020; Park et al. 2020; Park, Behrer, and Goodman 2021), while others study the impact of extreme temperatures on less challenging cognitive activities, with a focus on children and young adults (Garg, Jagnani, and Taraz 2020; Graff Zivin, Hsiang, and Neidell 2018). It remains unclear whether these findings hold true for more general population in low-stakes cognitive activities.

Our paper offers among the first evidence on transitory exposure to heat waves and low-stakes cognitive activities, leveraging a nationally representative longitudinal survey in China that includes almost all age cohorts, and matching with weather data according to the exact time and geographic location of the cognitive tests. Exploiting variations in exposure to extreme temperatures for the same individuals over eight years (2010-2018), we show that exposure to heat waves impedes performance in math tests. Specifically, exposure to a mean temperature above 32 °C on the test date, relative to a day in the 22– 24 °C range, leads to declined math test scores by 0.088 SDs (equivalent to 0.30 years of education). The effect is more pronounced for less educated older adults. We also observe salient cumulative effects of heat waves on both verbal and math tests. The analysis reveals that spending ten additional days in the year prior to the test with a daily average temperature above 28 °C, relative to a day in the 12–16 °C range, reduces verbal test scores by 0.142 (0.013 SDs) and math test scores by 0.098 (0.016 SDs), respectively.

This paper further provides suggestive evidence of adaptation to hot weather. Using data on residential air conditioning (AC) ownership at the household level, we show that AC offsets the detrimental effects of hot days (>32 °C) on cognition by 49.7 percent. People who live in hotter regions are less vulnerable to high temperatures.

We contribute to the literature on several fronts. First, including all groups above age 10 in low-stakes cognitive tests, we make the first attempt to identify heterogenous sensitivity to heat waves by age. Existing studies, however, mainly focus on the young children (Garg et al. 2020; Graff Zivin et al. 2018). Park et al. (2021) examine the age gradient of exposure to hot school days between students in elementary schools and those in middle schools. Our findings indicate that, while taking math tests, the impact of a day with a mean temperature above 32 °C, relative to a day in the 22-24 °C range, is on average 1.4 times as large on the elderly as on middle-aged people. People in old age are often faced with important choice making regarding retirement timing, pension premium, medical treatment, health insurance purchasing, other large spending, etc. Their more pronounced effects may disrupt cognitive ability to make such critical decisions. Cognitive decline or impairment in the elderly are also risk factors of Alzheimer’s disease, one of the costliest diseases, and other forms of dementia.

Second, with detailed information on individual-level residential AC status, we accurately assess the role of residential AC in the linkage between extreme temperatures and cognitive performance. Previous studies either rely on aggregated residential AC penetration data (e.g., Park et al. 2020) or impute probability of AC ownership based on social survey data (e.g., Graff Zivin et al. 2018). Our estimates indicate that adoption of AC at home offsets the negative effects of hot days (>32 °C) on cognition by 49.7 percent. These findings accounting for avoidance behaviors and defensive investments suggest substantial scope for adaptation to future climate change. In line with recent work exploring the extent to which AC mitigates the harmful effects of heat waves on mortality and labor productivity (Barreca et al. 2016; Behrer and Park 2017; Deschênes and Greenstone 2011), our results provide insights into the potential offsetting effects of adaptive behaviors, which are expected to play a critical role in determining the ultimate impacts of a changing climate.

Third, we are among the first to estimate the transitory impact of exposure to heat waves in a developing country setting and the benefits of residential AC. Garg et al. (2020) offer another evaluation of the transitory effect in an agrarian context in a single state of India, though the role of residential AC is not assessed. Penetration rates of residential AC differ vastly between developed countries and developing countries. For example, survey evidence suggests that while 90% of US households have some form of AC, only 34% and 13% of households in China and Mexico, respectively, own AC (Park et al. 2021). Given that the effects of climatic shocks on health-related outcomes vary substantially by socioeconomic status (Isen et al. 2017; Park et al. 2021), and that defensive investments such as AC can be effective in attenuating the impacts (Barreca et al. 2016; Behrer and Park 2017; Park et al. 2020), it is important to verify the external validity of evidence from high-income countries.

Finally, our findings also shed light on the various consequences of heat waves. Besides raising the disease burden (Deschênes and Greenstone 2011; Huang et al. 2012; Karlsson and Ziebarth 2018), increasing the risk of mental illness (Obradovich et al. 2018) and suicide rates (Burke et al. 2018), reducing labor supply (Deschenes 2014; Graff Zivin and Neidell 2014) as well as agricultural income and nutrition (Deschênes and Greenstone 2007; Shah and Steinberg 2017), we show that heat waves may impair intelligence, which depletes human capital, an important engine of economic growth. The total economic and social costs of heat waves are larger than previously thought, if we take the toll on intelligence into account.

The rest of the paper is organized as follows. Section 2 describes data sources. Section 3 discusses the empirical strategy. Section 4 reports our findings, including baseline results, adaptation and heterogeneous effects, as well as robustness checks. Finally, section 5 concludes.

## 2. Data

### 2.1. Cognition data

Data on cognitive tests are obtained from the China Family Panel Studies (CFPS), a nationally representative biennial longitudinal survey of Chinese families and individuals. CFPS is funded by Peking University and carried out by the university’s Institute of Social Science Survey.^1^ CFPS includes questions on a wide range of topics for families and individuals, including family dynamics and relationships, economic activities, health status, subjective well-being, and cognitive abilities.

The waves 2010, 2014 and 2018 of CFPS contain the same cognitive ability module, i.e., 24 standardized mathematics questions and 34 word-recognition questions. All these questions are obtained from standard textbooks and are sorted in ascending order of difficulty. The starting question depends on the respondent’s education level.^2^ The test ends when the individual incorrectly answers three questions in succession. The final test score is defined as the rank of the hardest question a respondent can answer correctly. If the respondent fails to answer any questions, the score is assigned as the rank of the starting question minus one. For example, a respondent with middle school education begins with the 9th question in the verbal test. If the hardest question one can correctly answer is the 14th question, then the verbal test scores would be 14. However, if one fails the 9th, 10th, and 11th questions consecutively, the verbal test score would be 8. Since the respondents did not know the testing rules prior to the interviews, there should be no incentive to manipulate test performance on purpose.

CFPS is suitable for our study for several reasons. First, the survey includes several standardized cognitive tests. Second, the survey embodies information on residential AC ownership, allowing us to study the adaptation behavior. Third, exact information about the geographic locations and test dates for all respondents is available, enabling us to precisely match individual test scores in the survey with local weather data. Further, the longitudinal data allow us to remove unobserved individual factors that may bias estimates. Finally, because the cognitive tests are administered to all age cohorts older than 10, we can study the effects of heat waves across age groups.

### 2.2. Weather data

The weather data are provided by the China National Meteorological Data Service Center (CMDC) under the National Meteorological Information Center of China. The dataset contains daily weather records of 824 monitoring stations along with their longitudes and latitudes in China. The key variable for our analysis was the daily mean temperature. Other weather controls include precipitation, wind speed, sunshine duration, and relative humidity. To merge the survey data with weather readings, we calculate the weighted average values of all the monitoring stations within a 100 km radius of each county centroid in CFPS, where the weights are equal to the inverse distance between stations and the county centroids.^3^ The spatial distribution of weather stations is displayed in Figure A1.^4^ The weather stations are evenly distributed in China and could be well matched with CFPS.

As studies show that air pollution is associated with bad performance in cognitive tests (Ebenstein, Lavy, and Roth 2016; Zhang, Chen, and Zhang 2018), we also add air quality as one of the control variables. Data on air quality are collected from the air quality report published by the Chinese Ministry of Ecology and Environment (MEE). The report includes 86 major cities in 2000 and covers most of the cities in China since 2014. Air quality is measured using the air pollution index (API), which ranges from 0 to 500, with larger values indicating worse air quality.^5^ We match each CFPS county to the nearest API reporting city within 100 km according to the distance between the county centroid and the city boundaries. All the key variables and their summary statistics are described in Table 1.

**Table 1.** Summary statistics.

| **Variable** | **Mean** | **Std. Dev.** |
| --- | --- | --- |
| **CFPS data** |  |  |
| verbal test scores | 18.284 | 10.553 |
| math test scores | 10.050 | 6.307 |
| gender | 0.479 | 0.500 |
| age | 46.508 | 17.458 |
| years of education | 7.422 | 4.585 |
| log household per capita income | 9.040 | 1.191 |
| **Temperature** |  |  |
| the **indicators** on the test date |  |  |
| <12 °C | 0.039 | 0.192 |
| 12-14 °C | 0.013 | 0.115 |
| 14-16 °C | 0.017 | 0.131 |
| 16-18 °C | 0.027 | 0.163 |
| 18-20 °C | 0.050 | 0.219 |
| 20-22 °C | 0.082 | 0.274 |
| 22-24 °C (*reference group*) | 0.120 | 0.324 |
| 24-26 °C | 0.156 | 0.363 |
| 26-28 °C | 0.173 | 0.378 |
| 28-30 °C | 0.197 | 0.398 |
| 30-32 °C | 0.110 | 0.313 |
| >32 °C | 0.016 | 0.125 |
| **Environmental controls** |  |  |
| API | 66.139 | 35.389 |
| precipitation, *mm* | 4.234 | 11.666 |
| wind speed, *m/s* | 2.101 | 0.934 |
| sunshine duration, *hour* | 6.465 | 4.166 |
| relative humidity, % | 74.509 | 12.106 |

Figure A2 depicts the histogram of mean temperatures on the test dates in our sample. As most of the interviews were conducted in July and August when college students were employed as numerators (Figure A3), the distribution is skewed to higher temperatures, with the mean being 24.74 °C. Following the general practice in the latest literature (Cho 2017; Graff Zivin et al. 2020, 2018), we use a series of indicators of 2 °C bin to allow for substantial flexibility and nonlinear relationships between the cognitive performance and temperature exposure. Specifically, we divide the temperature spectrum into 12 bins, with the lowest bin including all temperatures below 12 °C and the highest bin including all temperatures above 32 °C due to data sparseness at the extremities of the distribution. Figure 1 plots the percentage of days that fall into each bin, with 11.96 percent for the 22– 24 °C range, 19.71 for the 28-30 °C bin, and 1.59 for the greater than 32°C bin.

**Figure 1.**
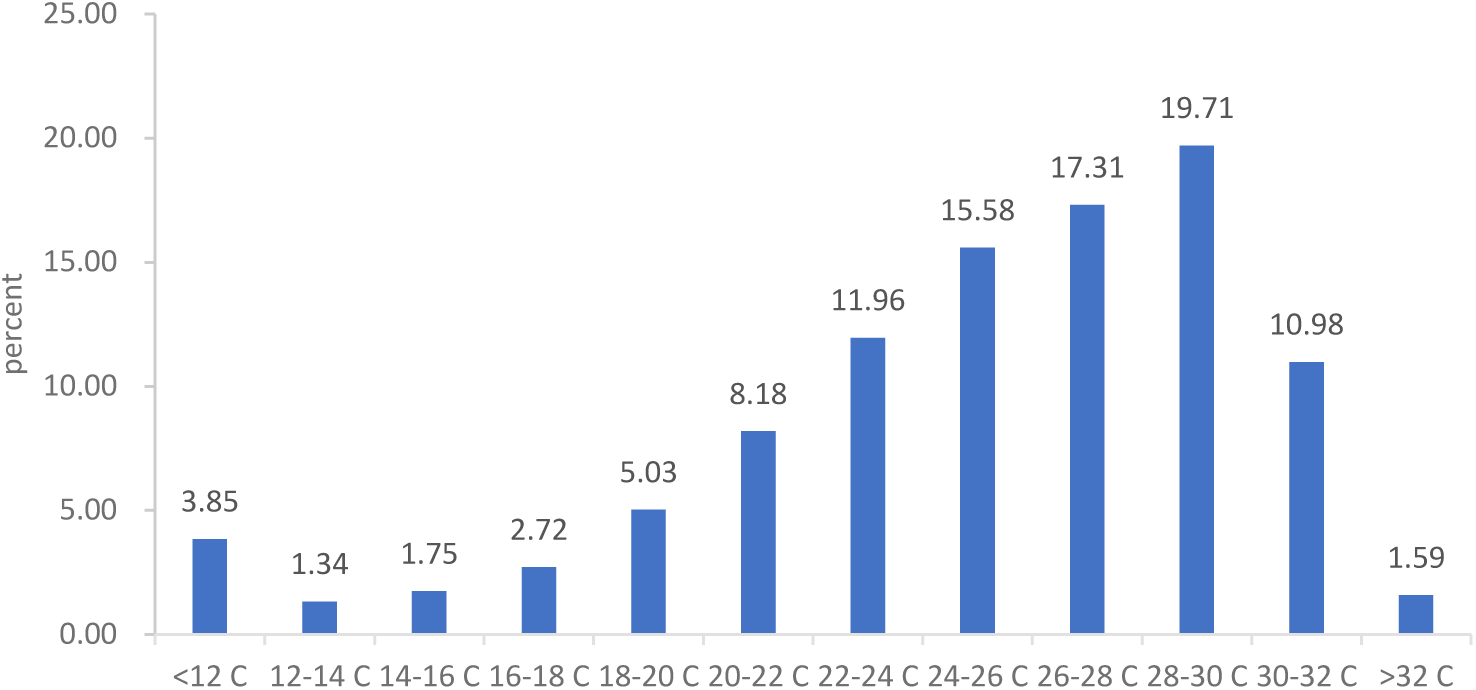
Distribution of daily mean temperature on the test date.

CFPS surveyed a panel of 49,652 individual respondents over age 10 in 2010, 2014 and 2018, for a total of 96,990 observations. Of these observations, 1,728 are missing values for test dates or locations. Among the remaining 95,262 observations, 71,776 observations could be matched to weather and API data. The matching rate of 75.3% (71,776 out of 95,262) is within a reasonable range compared with other studies (Levinson 2012). Due to some missing values for other control variables, the final dataset used in this study includes 70,687 observations.

## 3. Empirical strategy

Our baseline econometric specification is as follows:

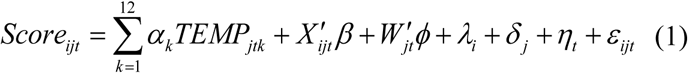

The dependent variable *Score*_*ijt*_ is the cognition test scores of respondent *i* in county *j* at date *t*. The two cognitive test scores we test in this paper are verbal test scores and math test scores. The key variables of interest *TEMP*_*jtk*_ are a series of indicators for whether the mean temperature falls into temperature bin *k* (from 1 to 12) on the test date *t* in county *j*. We deploy 12 bins, i.e., lower than 12 °C bin, higher than 32 °C bin, and ten 2 °C-wide bins in between. We set the 22–24 °C temperature bin as the reference group as it is associated with the highest cognitive test scores. The vector *X*_*ijt*_ represents demographic correlates, including age and its square term. We also control for a vector of contemporaneous air quality and weather conditions *W*_*jt*_, involving API, precipitation, wind speed, sunshine duration, and relative humidity in quadratic forms. *λ*_*i*_ denotes individual fixed effects. *δ*_*j*_ represents county fixed effects.^6^ *η*_*t*_ indicates year, month, and day of week fixed effects. *ε*_*ijt*_ is the error term. Standard errors are clustered at the county level.

By conditioning on the individual fixed effects and other sets of fixed effects listed above, the key parameters *α*_*k*_ are identified by making use of variations in exposure to temperatures for the same respondent in the three waves after controlling for seasonality and annual shocks. Due to the unpredictability of test dates and thus the random of temperature fluctuations, it is reasonable to assume that this variation is orthogonal to the unobserved determinants of cognitive test scores.

## 4. Results

### 4.1. Baseline results

Table A1 displays various specifications to our baseline results. Panel A corresponds to verbal tests, and Panel B is on math tests. The first column in each panel controls for temperature exposure, age and its square term, and individual fixed effects. We find a strong negative effect of exposure to heat waves (>32 °C) on math test scores. The pattern continues to hold when county fixed effects and a full set of year, month, and day-of-week fixed effects are added in the second column of each panel. Our preferred specification, with rich environmental conditions (i.e., API, total precipitation, wind speed, sunshine duration, and relative humidity in quadratic forms) further included, is displayed in the third column of each panel.

Figure 2 plots the estimated results from the preferred specification in Columns (3) and (6) of Table A1. Figure 2A corresponds to verbal test scores, while Figure 2B refers to math test scores. Each figure reveals the estimated coefficients for 12 temperature bins (<12 °C, 12–14 °C, 14–16 °C, 16–18 °C, 18–20 °C, 20–22 °C, 22–24 °C, 24–26 °C, 26– 28 °C, 28–30 °C, 30–32 °C, and >32 °C) in equation (1), together with their 90% and 95% confidence intervals (CIs). The temperature bin left out is 22–24 °C. Therefore, the coefficients for each bin measure the changes in test scores when temperature falls into that bin relative to the reference bin.

**Figure 2.**
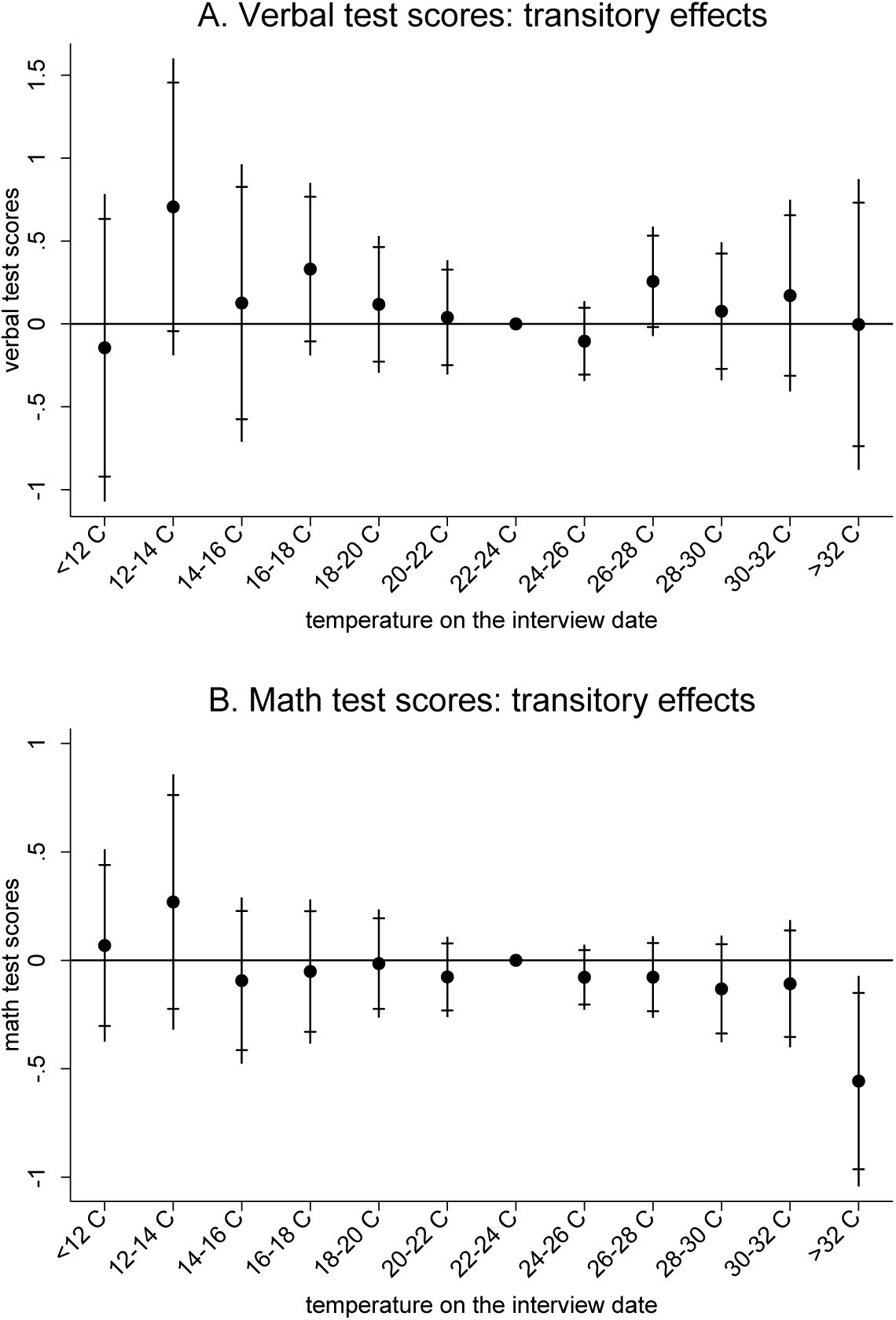
Transitory effects of temperature on cognitive test scores. Note: The figures plot the estimated coefficients on temperature bins based on the results in Columns (3) and (6) of Table A1. Both 90 (short caps) and 95 percent (long lines) confidence intervals are displayed. The left-out temperature bin is 22-24 °C. The coefficients can be interpreted as effects of a day in the corresponding temperature bin on cognitive test scores relative to the reference temperature category. Panel A refers to verbal test scores, while Panel B refers to math test scores.

As revealed in Figure 2A, there is no obvious association between temperature exposure and cognitive performance in verbal tests. All the coefficients on temperature bins are insignificant. Heat waves seem to have little effect on verbal test scores. Figure 2B further presents the estimated effect on math test scores. We find a non-linear relationship between temperature and cognitive performance in math tests, where both low and high temperatures are associated with declines in math test scores. When exposed to temperatures higher than 32 °C, the negative effect is significant at the 5% level. Specifically, a test day with a mean temperature above 32 °C, relative to a day in the 22– 24 °C range, leads to a reduction in math test scores by 0.557. To put this into context, note that the standard deviation (SD) of math test scores is 6.307. Therefore, respondents’ math test scores, on days with average temperatures above 32 °C, are on average 0.088 SDs lower than their scores on a day in the reference temperature bin (22–24 °C).

Our results by test subject are consistent with the literature in which the transitory effect of exposure to high temperatures is more often observed in math tests than in other subjects like word recognition and reading comprehension (Garg et al. 2020; ZGraff ivin et al. 2018; Park 2020). One potential explanation is that different regions of the brain perform distinct cognitive functions, and the regions responsible for solving math problems are more sensitive to extreme temperatures than the regions in charge of reading functions (Hocking et al. 2001). These differential effects in the short run across cognitive tasks also provide strong evidence for the presence of a physiological channel connecting temperature exposures to cognitive performance.

We further interpret our findings in two ways. First, we calculate years of education lost using the estimates, as cognition and educational attainment are highly correlated and intrinsically linked. Figure A4 plots average years of education on cognitive test scores and estimates the correlation coefficients between these two variables for verbal and math test scores, respectively. A one-point increase in verbal test scores corresponds to 0.318 years of education, while a one-point increase in math test scores is equivalent to 0.537 years of education. Calculated from Table A1, exposure to a mean temperature above 32 °C on the test date, relative to a day in the 22–24 °C range, leads to a sizable decrease in math test scores by 0.30 years of education.

Second, we estimate the effect size in terms of changes in test score percentile. Figure A5 plots the percentile of scores for verbal and math tests, respectively. Exposure to a mean temperature above 32 °C on the test date, relative to a day in the 22–24 °C range, leads to a sizable decline in math test scores, equivalent to moving from the 51.4th percentile in the math test distribution to the median (i.e., the 50th percentile).

### 4.2. Adaptation and heterogeneous effects

We study adaptation to heat waves in two dimensions. First, we split the sample according to the residential AC ownership. As shown in Figures 3A for verbal test scores and in Figures 3B for math test scores, the negative effect of exposure to heat waves is significant only for individuals taking math tests without AC. As reported in Columns (3)-(4) of Table A2, adoption of AC offsets some of the negative effects of hot days (>32 °C) on cognition. The effect size of heat waves on math test scores with AC is 49.7% smaller than that without AC.

**Figure 3.**
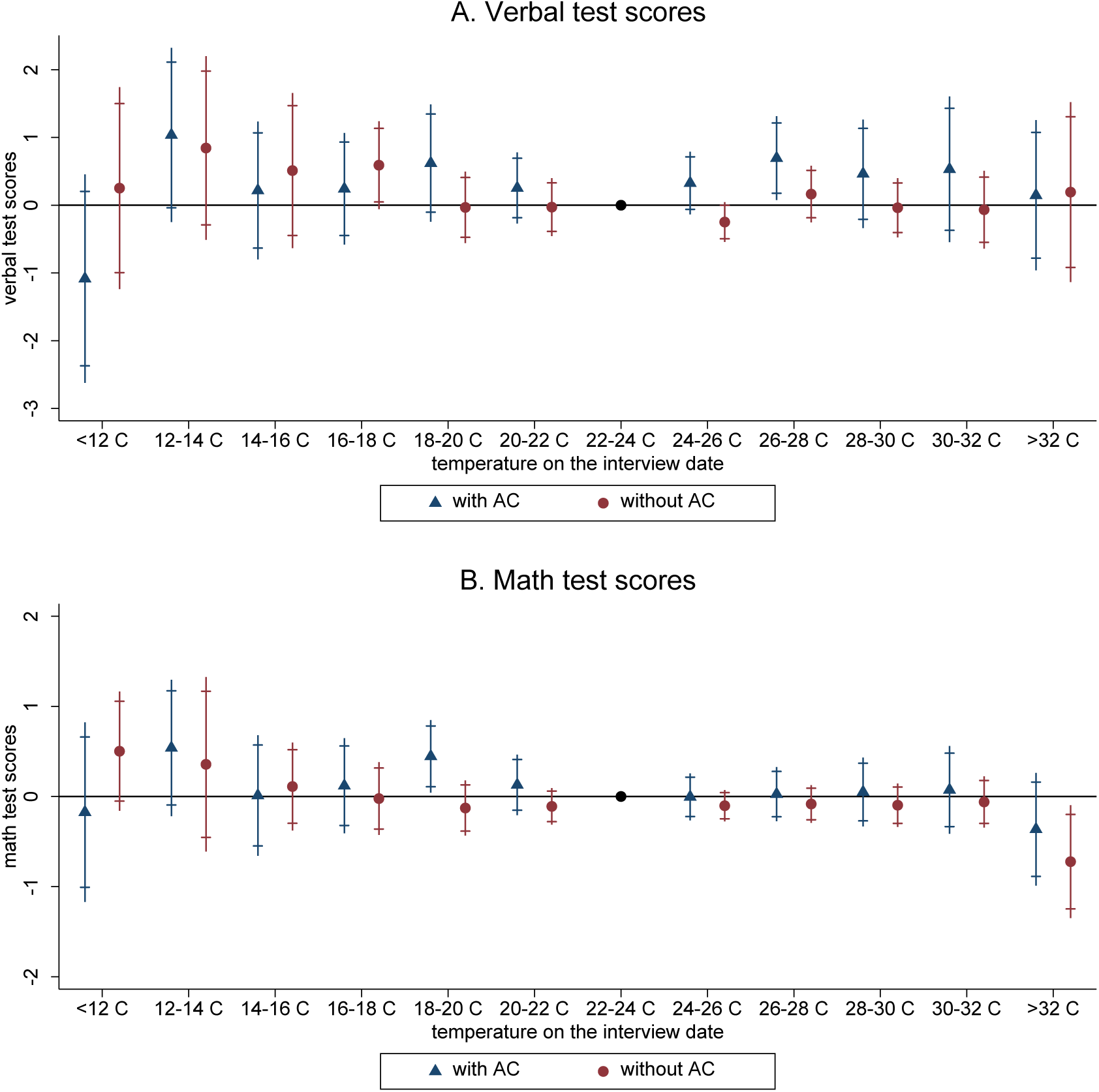
Transitory effects of temperature on cognitive test scores, by residential AC ownership. Note: The figures plot the estimated coefficients on temperature bins for the households with and without AC based on the results in Columns (1) through (4) of Table A2. Both 90 (short caps) and 95 percent (long lines) confidence intervals are displayed. The left-out temperature bin is 22-24 °C. The coefficients can be interpreted as effects of a day in the corresponding temperature bin on cognitive test scores relative to the reference temperature category. Panel A refers to verbal test scores, while Panel B refers to math test scores.

Second, we repeat the exercises for cooler and hotter regions of China, classified according to the median of each county’s average temperature in summer days (from June to August) during 2010-2018. The results are plotted in Figure 4, and the corresponding numerical results are displayed in Columns (5)-(8) of Table A2. As revealed in Figure 4, people living in cooler regions are more sensitive to heat waves than those living in hotter regions. In the cooler regions, math test scores on days with average temperature above 32 °C are on average 1.308 (0.207 SDs) lower than their scores on a day in the reference temperature bin (22–24 °C). By contrast, cold spells (<12 °C) are more harmful to the verbal test performance of respondents living in hotter regions than cooler regions. Our findings are consistent with the literature on adaptation behavior. For example, Cho (2017) shows that students in cities with relatively cool summers are affected more than students in cities with relatively hot summers. Behrer and Park (2017) find that very hot places in the US seem to better adapt to heat stress than cooler areas.

**Figure 4.**
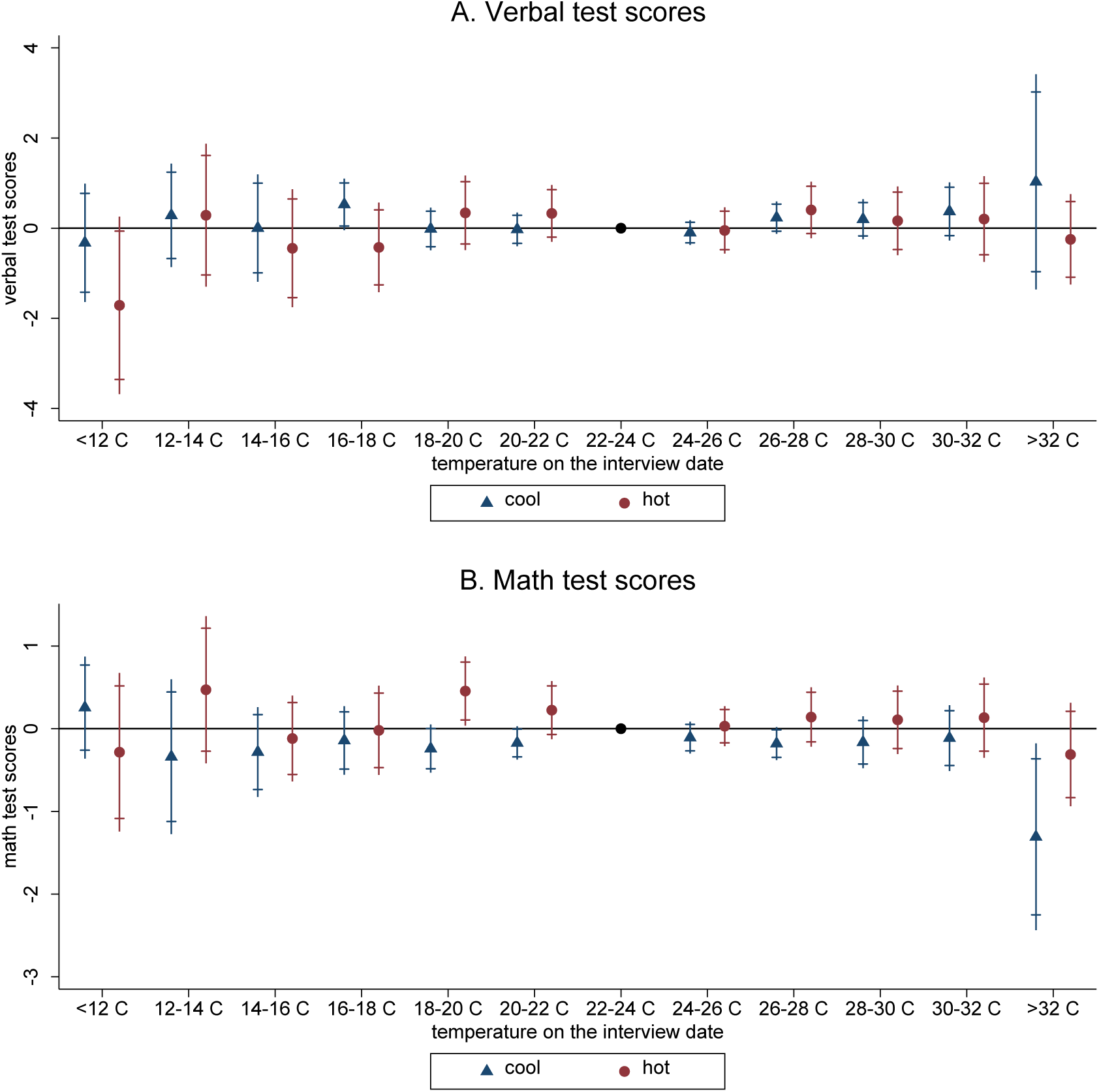
Transitory effects of temperature on cognitive test scores, by region. Note: The figures plot the estimated coefficients on temperature bins for the cool and hot regions based on the results in Columns (5) through (8) of Table A2. Both 90 (short caps) and 95 percent (long lines) confidence intervals are displayed. The left-out temperature bin is 22-24 °C. The coefficients can be interpreted as effects of a day in the corresponding temperature bin on cognitive test scores relative to the reference temperature category. Panel A refers to verbal test scores, while Panel B refers to math test scores.

We also examine the heterogeneous effect by gender, age and education level. Figure 5 firstly presents the estimated results by gender. When exposed to temperatures higher than 32 °C, the negative effect on math test scores is significant for both males and females. However, the effect is muted for cognitive performance in verbal tests. Women seem to be a little more vulnerable than men to heat waves, but the gender difference is weak. As reported in Columns (3)-(4) of Table A3, spending a day with a temperature above 32 °C, relative to a day in the 22–24 °C range, decreases males’ and females’ math test scores by 0.575 (0.091 SDs) and 0.608 (0.096 SDs), respectively.

**Figure 5.**
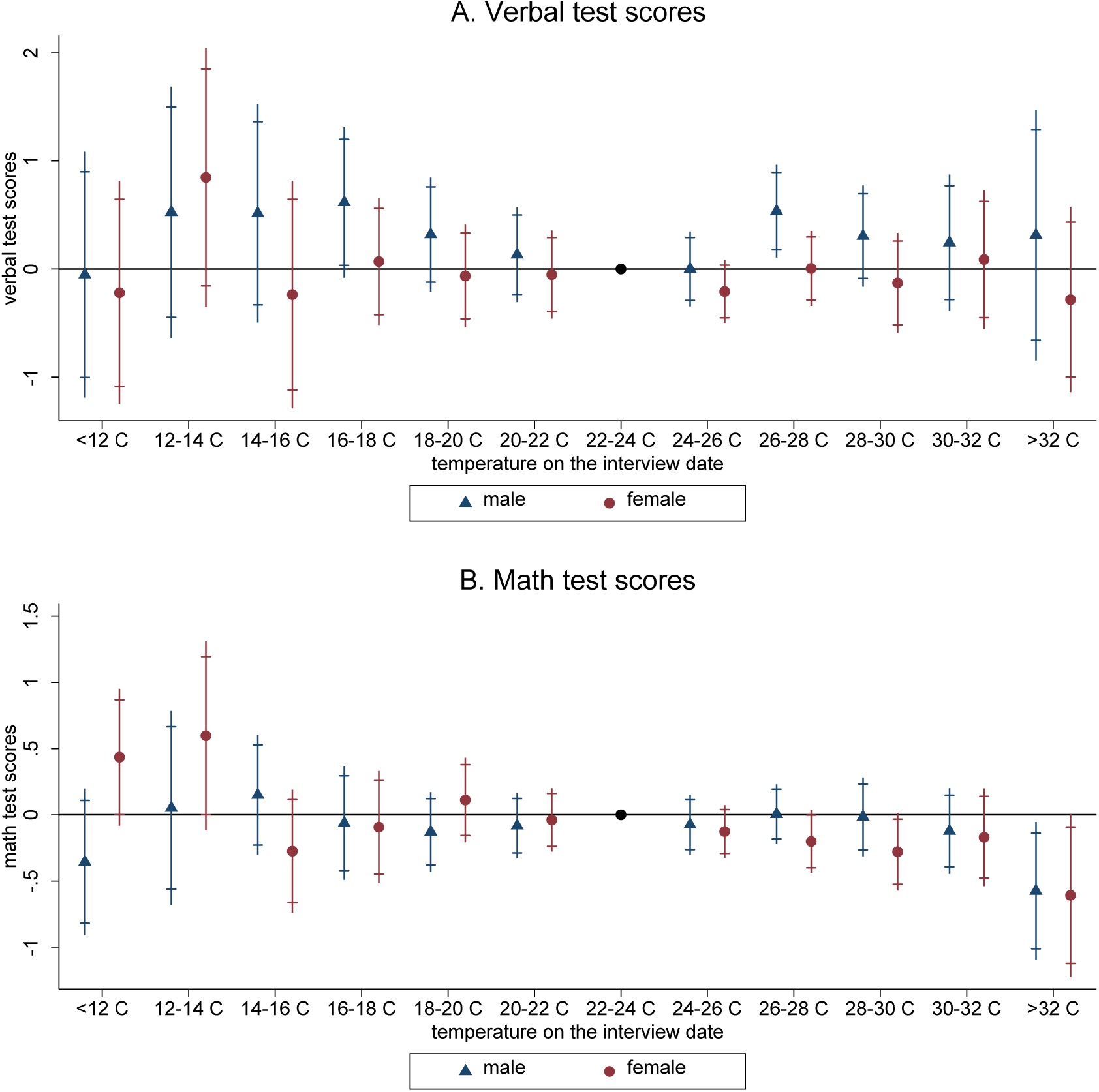
Transitory effects of temperature on cognitive test scores, by gender. Note: The figures plot the estimated coefficients on temperature bins for the male and female subsamples based on the results in Columns (1) through (4) of Table A3. Both 90 (short caps) and 95 percent (long lines) confidence intervals are displayed. The left-out temperature bin is 22-24 °C. The coefficients can be interpreted as effects of a day in the corresponding temperature bin on cognitive test scores relative to the reference temperature category. Panel A refers to verbal test scores, while Panel B refers to math test scores.

To explore whether the effects of heat waves on cognition differ across age cohorts, we divide the sample into three age groups (10-30, 31-59 and 60+). Figure 6 plots the estimates and CIs on the temperature bins for the three age cohorts, separately. Figure 6A displays the results for verbal tests, while Figure 6B presents the estimates for math tests. The numerical results are shown in Table A4. Compared to the younger cohort, heat waves are more harmful to middle-aged or older adults taking math tests, especially among older adults. Specifically, a day with a mean temperature above 32 °C, relative to a day in the 22–24 °C range, is associated with a 0.564 (0.089 SDs) and 0.778 (0.123 SDs) decline in math test scores for the middle-aged (age 31-59) and the old people (age 60 or above), respectively.

**Figure 6.**
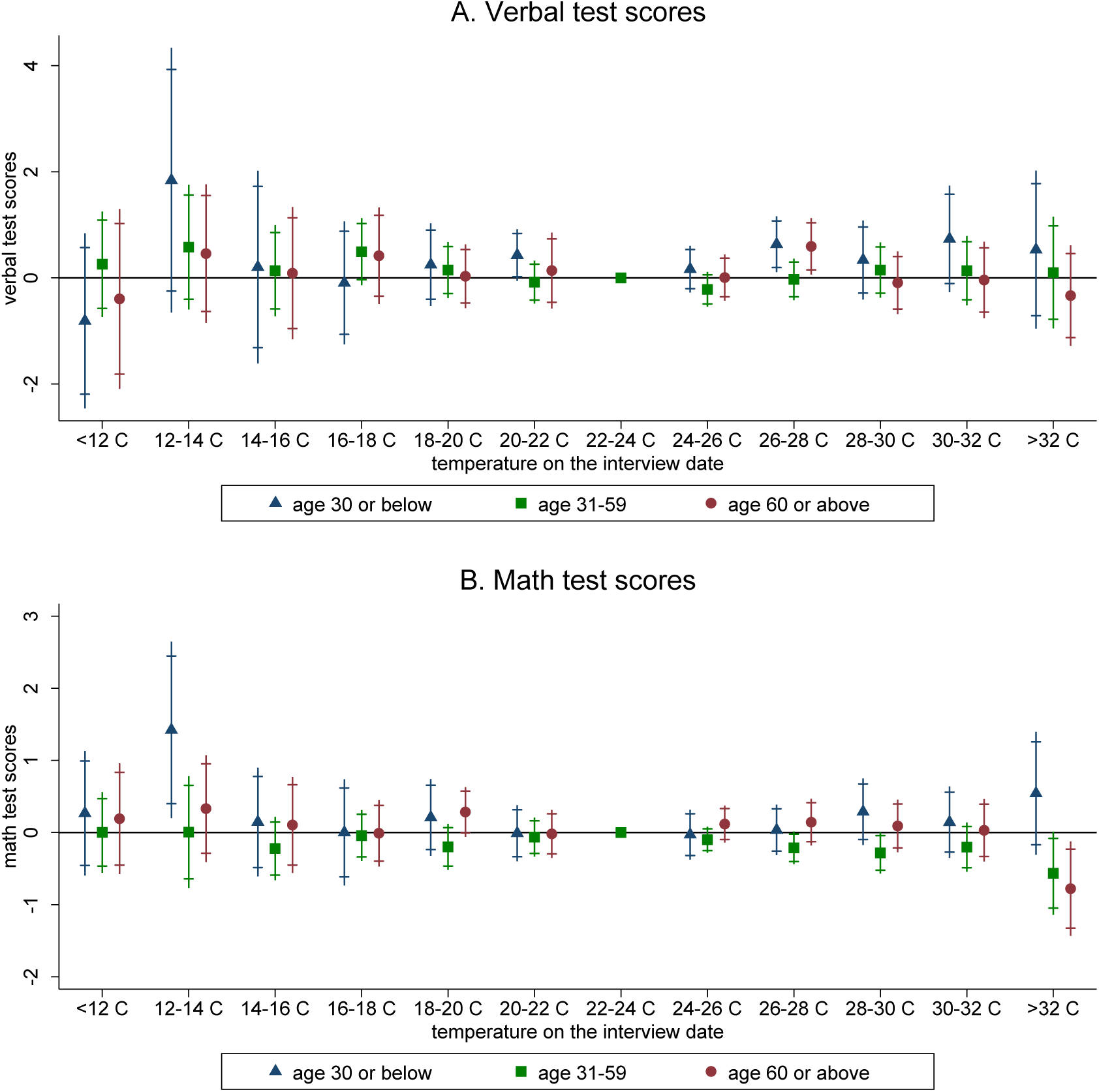
Transitory effects of temperature on cognitive test scores, by age cohort. Note: The figures plot the estimated coefficients on temperature bins for the age cohorts based on the results in Table A4. Both 90 (short caps) and 95 percent (long lines) confidence intervals are displayed. The left-out temperature bin is 22-24 °C. The coefficients can be interpreted as effects of a day in the corresponding temperature bin on cognitive test scores relative to the reference temperature category. Panel A refers to verbal test scores, while Panel B refers to math test scores.

Finally, education level may have a significant effect on response to extreme high temperatures. Dividing the whole sample into two subgroups at 12 years of education, Figure 7B shows that heat waves impose a significant effect on math tests of less educated people. The numerical results in Table A3 indicate that a day with a mean temperature above 32 °C, relative to a day in the 22–24 °C range, leads to a reduction in math test scores by 0.582 (0.092 SDs) for respondents who received 0-12 years of education.

**Figure 7.**
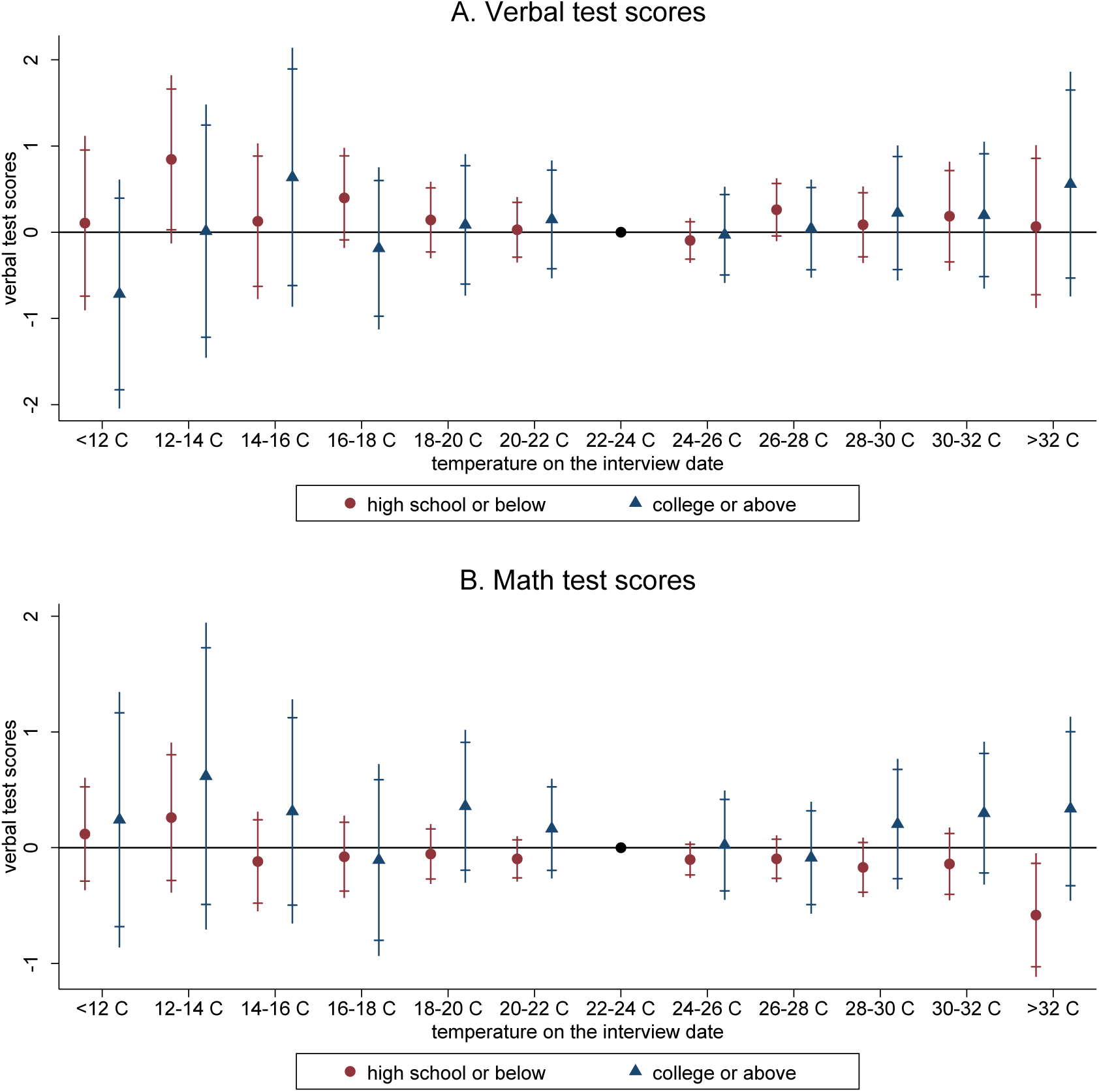
Transitory effects of temperature on cognitive test scores, by education level. Note: The figures plot the estimated coefficients on temperature bins for the less educated (high school or below) and educated (college or above) based on the results in Columns (5) through (8) of Table A3. Both 90 (short caps) and 95 percent (long lines) confidence intervals are displayed. The left-out temperature bin is 22-24 °C. The coefficients can be interpreted as effects of a day in the corresponding temperature bin on cognitive test scores relative to the reference temperature category. Panel A refers to verbal test scores, while Panel B refers to math test scores.

### 4.3. Robustness checks

We conduct a placebo test to address the concern over potential omitted variables. Following a common strategy in the literature (Cho 2017), we examine the effect of extreme temperature on the same day next year on cognitive test scores. If unobserved factors are correlated with both the time trend of extreme temperature and the outcome variables, we would find similar effects when replacing the current exposure with later ones. Columns (1)-(2) of Table 2 display the estimated coefficients from regressions of verbal and math test scores on temperature bins one year after the interview. None of the coefficients is statistically different from zero, which largely dismisses the concern over omitted variables.

**Table 2.** Robustness checks.

| Dependent variable | Placebo test |  | Ruling out other behavioral channels |  | Using non-migrants only |  | Excluding ozone dominated days |  |
| --- | --- | --- | --- | --- | --- | --- | --- | --- |
|  | verbal | math | cooperation | impatience | verbal | math | verbal | math |
|  | (1) | (2) | (3) | (4) | (5) | (6) | (7) | (8) |
| temperature bins |  |  |  |  |  |  |  |  |
| <12 °C | -0.012<br>(0.658) | 0.051<br>(0.318) | -0.302<br>(0.202) | -0.199<br>(0.392) | -0.091<br>(0.479) | 0.074<br>(0.230) | -0.270<br>(0.505) | 0.064<br>(0.258) |
| 12-14 °C | -0.654<br>(0.714) | -0.187<br>(0.325) | -0.324<br>(0.201) | 0.051<br>(0.330) | 0.636<br>(0.460) | 0.257<br>(0.305) | 0.401<br>(0.509) | 0.337<br>(0.350) |
| 14-16 °C | -0.087<br>(0.412) | -0.126<br>(0.201) | -0.028<br>(0.148) | -0.187<br>(0.364) | 0.067<br>(0.428) | -0.119<br>(0.200) | 0.154<br>(0.467) | -0.070<br>(0.211) |
| 16-18 °C | -0.086<br>(0.290) | 0.189<br>(0.149) | -0.057<br>(0.128) | -0.451<br>(0.304) | 0.299<br>(0.268) | -0.071<br>(0.172) | 0.168<br>(0.334) | -0.046<br>(0.187) |
| 18-20 °C | -0.050<br>(0.217) | -0.078<br>(0.104) | -0.090<br>(0.111) | -0.269<br>(0.202) | 0.129<br>(0.210) | -0.022<br>(0.126) | 0.040<br>(0.259) | -0.014<br>(0.139) |
| 20-22 °C | -0.062<br>(0.181) | -0.070<br>(0.082) | -0.060<br>(0.078) | 0.042<br>(0.173) | 0.051<br>(0.175) | -0.069<br>(0.095) | -0.102<br>(0.212) | -0.121<br>(0.100) |
| 22-24 °C |  |  |  |  |  |  |  |  |
| 24-26 °C | 0.028<br>(0.148) | 0.073<br>(0.074) | -0.094*<br>(0.051) | 0.117<br>(0.139) | -0.110<br>(0.124) | -0.082<br>(0.077) | -0.051<br>(0.165) | -0.065<br>(0.097) |
| 26-28 °C | 0.002<br>(0.182) | 0.056<br>(0.091) | -0.107<br>(0.092) | 0.067<br>(0.168) | 0.281*<br>(0.166) | -0.068<br>(0.095) | 0.244<br>(0.215) | -0.135<br>(0.116) |
| 28-30 °C | 0.205<br>(0.199) | 0.048<br>(0.105) | -0.051<br>(0.095) | 0.134<br>(0.183) | 0.088<br>(0.213) | -0.123<br>(0.126) | -0.046<br>(0.278) | -0.222<br>(0.156) |
| 30-32 °C | 0.284<br>(0.247) | -0.028<br>(0.123) | -0.158<br>(0.116) | 0.178<br>(0.223) | 0.182<br>(0.300) | -0.106<br>(0.152) | 0.202<br>(0.433) | 0.012<br>(0.190) |
| >32 °C | -0.143<br>(0.351) | 0.091<br>(0.141) | -0.255<br>(0.203) | 0.025<br>(0.321) | 0.001<br>(0.460) | -0.558**<br>(0.252) | -0.440<br>(0.422) | -0.768***<br>(0.273) |
| Observations | 70,445 | 70,445 | 47,052 | 48,530 | 68,543 | 68,543 | 54,673 | 54,673 |
| Adjusted R-squared | 0.775 | 0.785 | 0.113 | 0.069 | 0.774 | 0.782 | 0.778 | 0.810 |

Note: The placebo test is conducted using temperature exposure the same day next year. The left-out temperature bin is 22-24 °C. The coefficients can be interpreted as effects of a day in the corresponding temperature bin on cognitive test scores relative to the reference temperature category. All the regressions include individual fixed effects, county fixed effects, year, month, and day-of-week fixed effects. Demographic controls include age and its square term. Environmental controls include air pollution index (API), total precipitation, wind speed, sunshine duration, and relative humidity in quadratic forms. Robust standard errors, clustered at the county level, are presented in parentheses. \* 10% significance level; \*\* 5% significance level; \*\*\* 1% significance level.

The transitory effect of heat waves on cognitive performance may be driven by behavioral change. People may become less cooperative or more impatient when exposed to extreme high temperatures, thereby reducing their cognitive test scores. CFPS includes evaluation of interviewees’ level of cooperation (waves 2010 and 2014) and impatience (waves 2014 and 2018), as rated by the interviewers. The ratings for cooperation and impatience are both scaled from 1 (low) to 7 (high). We explore the effect of exposure to extreme temperatures on cooperation and impatience using a specification similar to equation (1). Table 2 reports the results for cooperation and impatience in Columns (3) and (4), respectively. The estimates indicate no significant association between extreme high temperature (>32 °C) and respondents’ cooperation or impatience, ruling out the behavioral channel. As described earlier, differences in the heat sensitivity of the brain regions are more plausible channels through which heat waves may disrupt cognition.

We have so far used daily mean temperature in the analyses. In Figure 8, we instead use the daily maximum temperature. We divide the daily maximum temperature into 12 bins, i.e., <18 °C, 18-20 °C, 20-22 °C, 22-24 °C, 24-26 °C, 26-28 °C, 28-30 °C, 30-32 °C, 32-34 °C, 34-36 °C, 36-38 °C, and >38 °C. Figure A6 shows their distribution in our sample. As reported in Figure 8, the similar effects persist. Specifically, individuals’ math test scores are on average 0.555 (0.088 SDs) lower on a day with a maximum temperature above 38 °C than with a maximum temperature of 28–30 °C.

**Figure 8.**
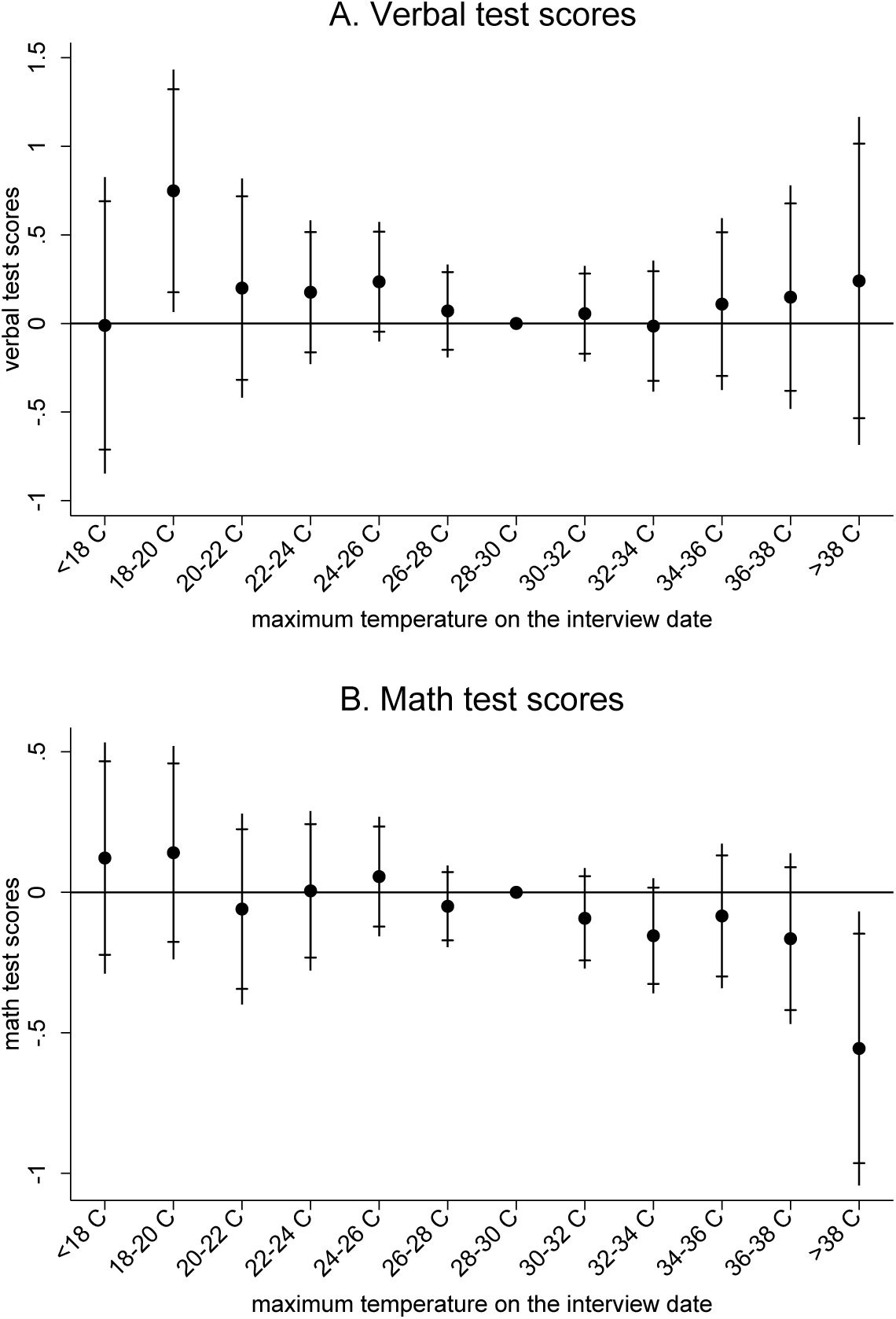
Robustness check –maximum temperature bins. Note: The figures plot the estimated coefficients on maximum temperature bins “<18 °C, 18-20 °C, 20-22 °C, 22-24 °C, 24-26 °C, 26-28 °C, 28-30 °C, 30-32 °C, 32-34 °C, 34-36 °C, 36-38 °C, and >38 °C” with 90% and 95% confidence intervals. The left-out temperature bin is 28-30 °C. The coefficients can be interpreted as effects of a day in the corresponding temperature bin on cognitive test scores relative to the reference temperature category. Panel A refers to verbal test scores, while Panel B refers to math test scores.

The results are also robust to a range of alternative specifications. Columns (5)-(6) of Table 2 indicates that migration, and thus location sorting, is unlikely to significantly bias our estimates. Columns (7)-(8) of Table 2 reveals that our findings still hold after we exclude ozone dominated days, during which ozone may further interact with heat waves to impair cognition.

Finally, we provide some evidence on the *cumulative* effect of exposure to heat waves on cognitive performance. Following the specification in Park et al. (2020), who studies the effect of hot school days in the 365 days prior to the test on students’ performance in high-stakes exams, we calculate the number of days falling in each temperature bin (<-4 °C, -4-0 °C, 0-4 °C, 4-8 °C, 8-12 °C, 12-16 °C, 16-20 °C, 20-24 °C, 24-28 °C, and >28 °C) in the past year before the interview. Figure A7 shows the distribution of the cumulative exposure and Figure 9 plots the estimated coefficients associated with each temperature bin for verbal and math test scores with 90% and 95% CIs. The temperature bin left out is 12–16 °C. The coefficients can be interpreted as effects of an additional day in the corresponding temperature bin on cognitive test scores relative to the reference temperature category. Different from transitory effect, results indicate significant effects of cumulative exposure to extreme high temperatures (>28 °C) on both verbal and math test scores. Cold days (below -4 °C) appear to have impact on cognitive performance in math tests. The estimates reveal that spending ten additional days in the year prior to the test with a daily average temperature above 28 °C, relative to a day in the 12–16 °C range, reduces verbal test scores by 0.142 (0.013 SDs) and math test scores by 0.098 (0.016 SDs), respectively.

**Figure 9.**
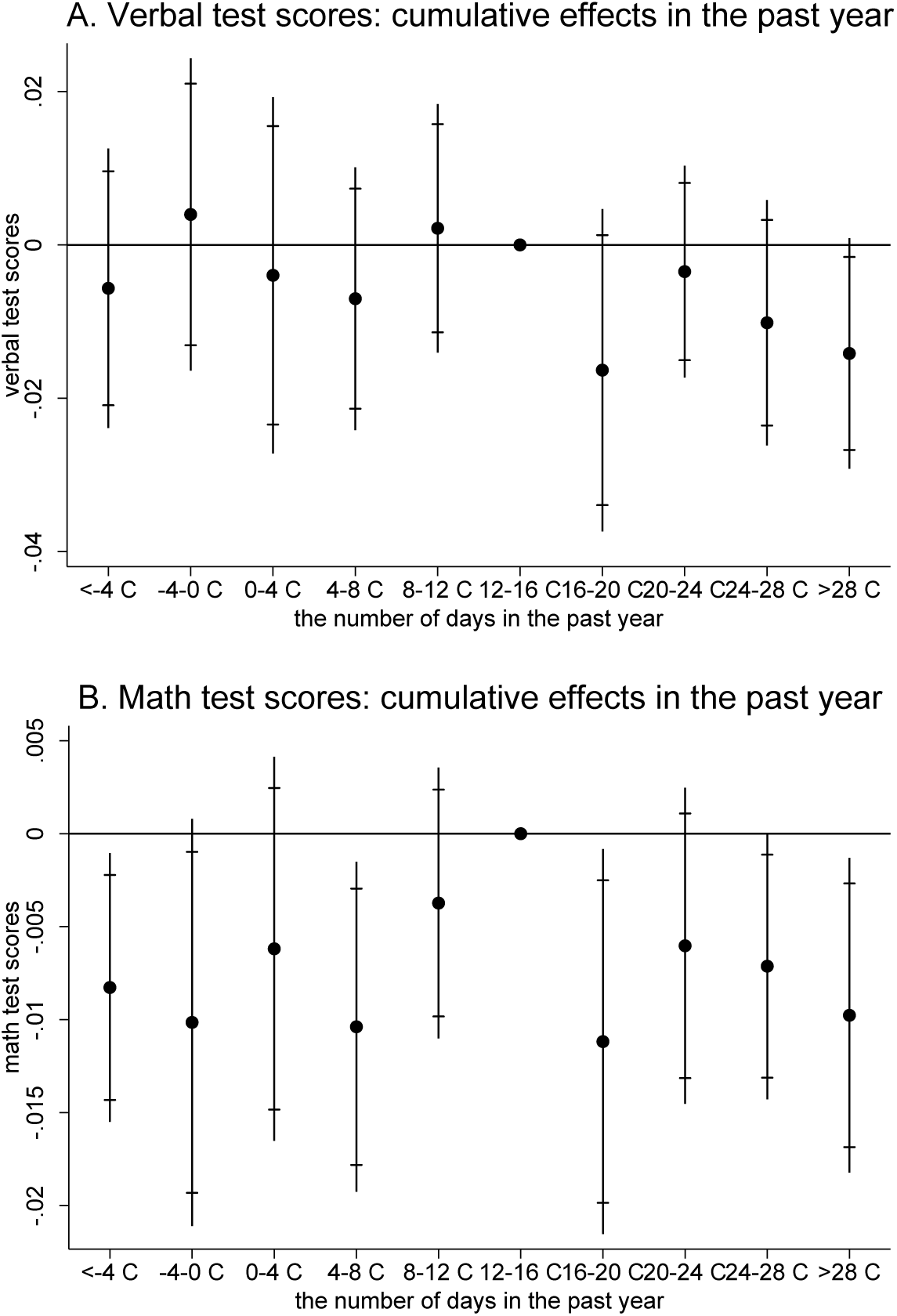
Cumulative effects of temperature on cognitive test scores. Note: The figures plot the estimated coefficients on the number of days in each temperature bin “<-4 °C, -4-0 °C, 0-4 °C, 4-8 °C, 8-12 °C, 12-16 °C, 16-20 °C, 20-24 °C, 24-28 °C, and >28 °C” during the past year. The left-out temperature bin is 12-16 °C. Both 90 (short caps) and 95 percent (long lines) confidence intervals are displayed. The coefficients can be interpreted as effects of an additional day in the corresponding temperature bin on cognitive test scores relative to the reference temperature category. Panel A refers to verbal test scores, while Panel B refers to math test scores.

## 5. Conclusions

Matching a nationally representative longitudinal survey with weather data according to the exact date and geographic location in China, this study examines the effect of transitory exposure to heat waves on cognitive performance for people above age 10. Exploiting the longitudinal structure of CFPS and random fluctuations in weather across interviews, we identify the effect of temperatures in models with individual fixed effects. We find that exposure to a mean temperature above 32 °C on the test date, relative to a day in the 22–24 °C range, leads to a decline in math test scores by 0.088 SDs, which can be converted to 0.30 years of education or moving from the 51.4th percentile to the median in the math score distribution. The effect is more pronounced for older adults who are less educated. People living in hotter regions or having AC installed in their homes are less vulnerable to extreme high temperatures, indicating some adaptation. We provide the first evidence that residential AC could mitigate the harmful effect of heat waves on math test scores by 47.9%. The results survive a placebo test and a set of robustness checks.

Our findings highlight the distributional consequences of climate change. People in lower SES are more vulnerable to heat waves, and uneven access to avoidance options such as AC may exacerbate social inequities. Previous studies evaluating the welfare cost of climate change have neglected its potential effect on cognitive decline among older adults. As people often spend most of their wealth and make critical decisions in old age, the salient impact on the elderly suggests that a narrow focus on the young may underestimate the social costs of heat waves on cognition.

While this study mainly focuses transitory exposures to heat events, the impact is sizable. First, our results indicate that cognitive ability can be compromised even in temperatures (>32 °C) below commonly recognized hot days. Second, compared to previous work, cognitive tests in our study setting are close to our day-to-day, low-stakes cognitive activities. The salient effect in our setting suggests that the quality of routine decision-making in our daily lives is affected by heat waves. Third, although the effect of exposure to heat waves is transitory, more frequent heat waves associated with climate change will likely impede cognitive performance more in the future. Cognitive function is essential for daily decision making. Damages to cognitive performance caused by heat waves would compromise the quality of decision-making, generating inefficiencies and imposing additional costs on individual and social welfare. These hidden costs of climate change should be considered in climate policy assessments.

## Data Availability

All data produced in the present study are available upon reasonable request to the authors.

https://www.isss.pku.edu.cn/cfps/en/index.htm?CSRFT=Q9BL-UGK3-26PK-N7FS-QJW9-V1DT-5JX5-U7RI

## Acknowledgements

We acknowledge the Institute of Social Science Survey at Peking University for providing us with the CFPS data. The authors acknowledge helpful comments by participants and discussants at the various conferences, seminars and workshops.

## Funding information

Xi Chen acknowledges funding from the Yale Macmillan Center Faculty Research Fund, the US Federal PEPPER Center Scholar Award (P30AG021342), two NIH/National Institute on Aging Grants (R03AG048920 and K01AG053408). Xin Zhang acknowledges financial support from the National Natural Science Foundation of China (72003014).

## Conflict of interest

The authors declare that they have no conflict of interest.

## Ethics approval

The study was approved by the Institutional Review Board (IRB) at Peking University (Approval No: IRB00001052-14010). All participants gave informed consent in accordance with policies of the IRB at Peking University.

## Online Supplementary Appendix

**Figure A1.**
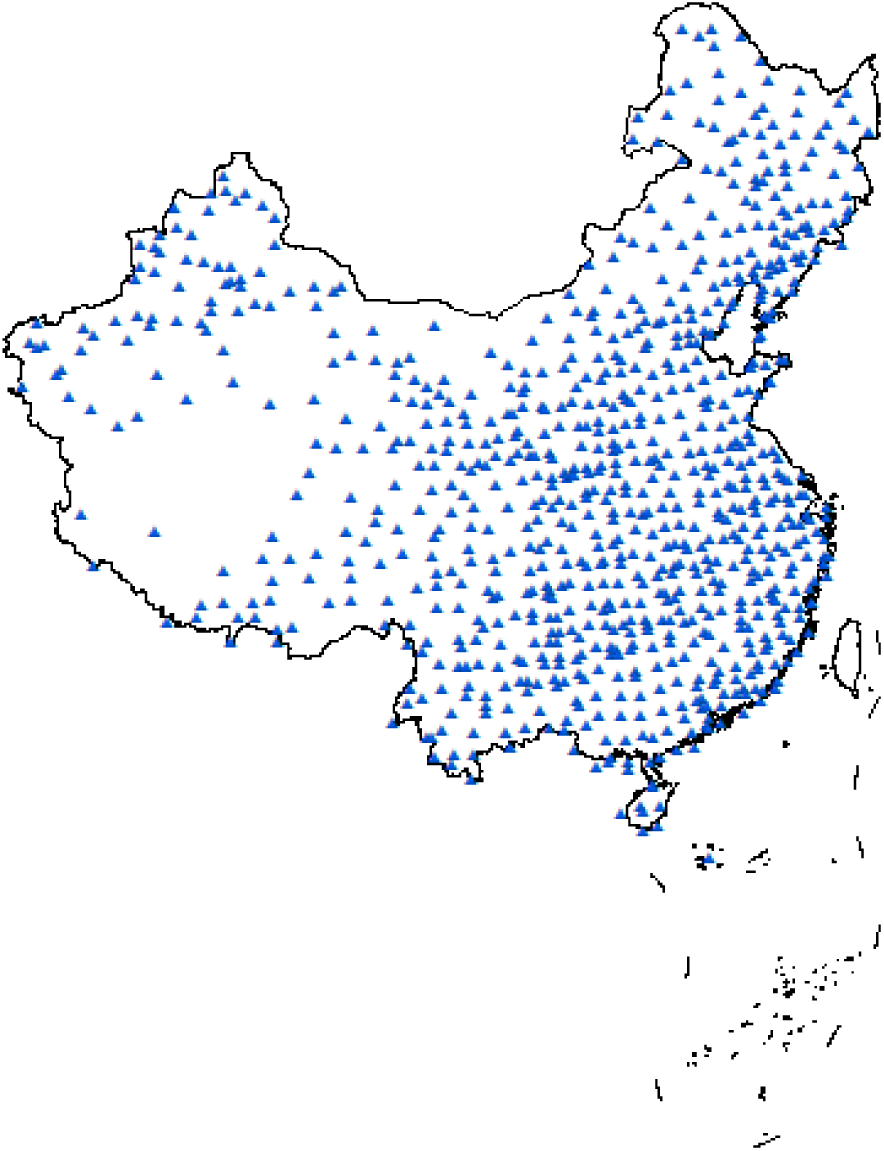
The distribution of weather stations. Note: This figure is plotted using ArcMap 10.3.1.

**Figure A2.**
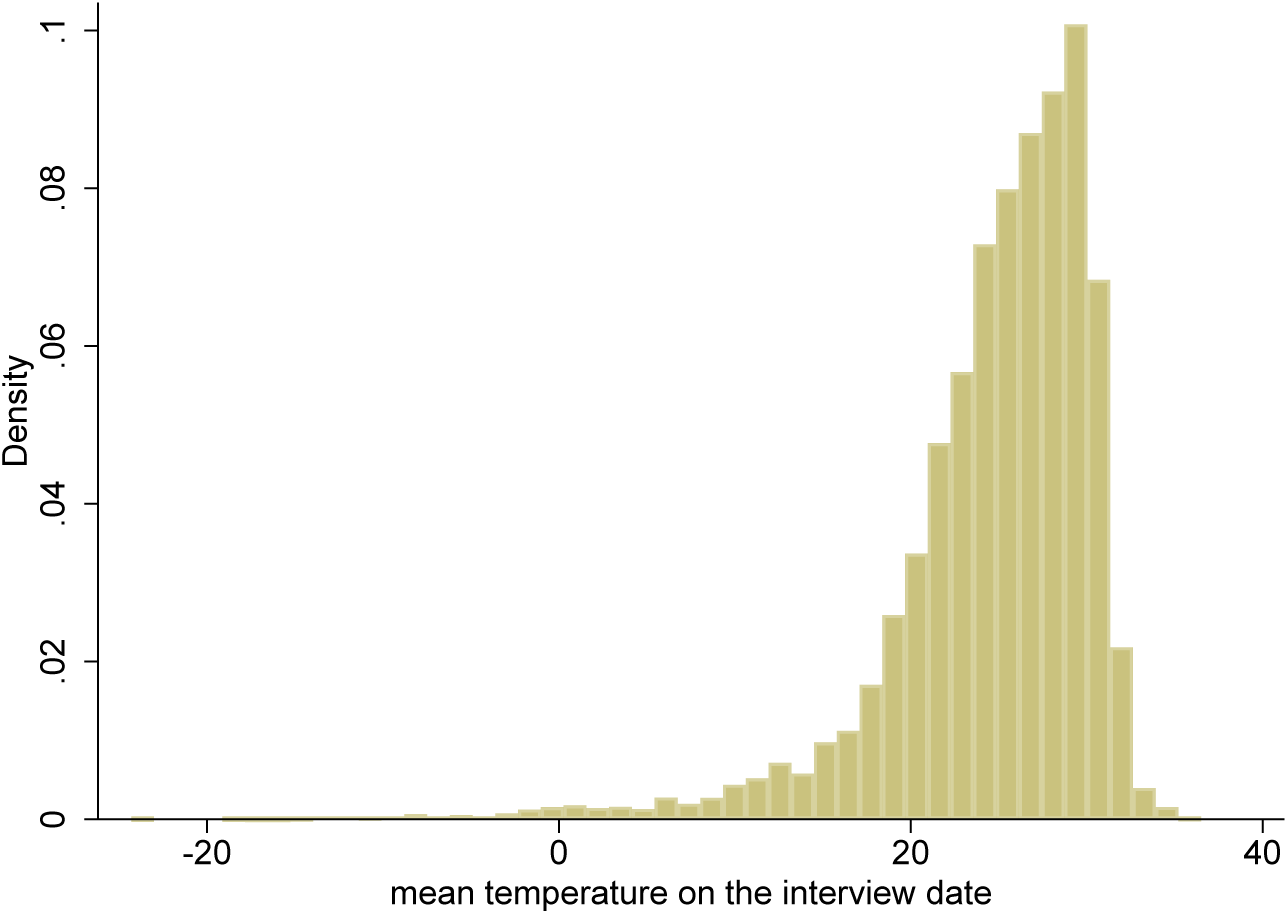
Histogram of mean temperature (°C) on the test date.

**Figure A3.**
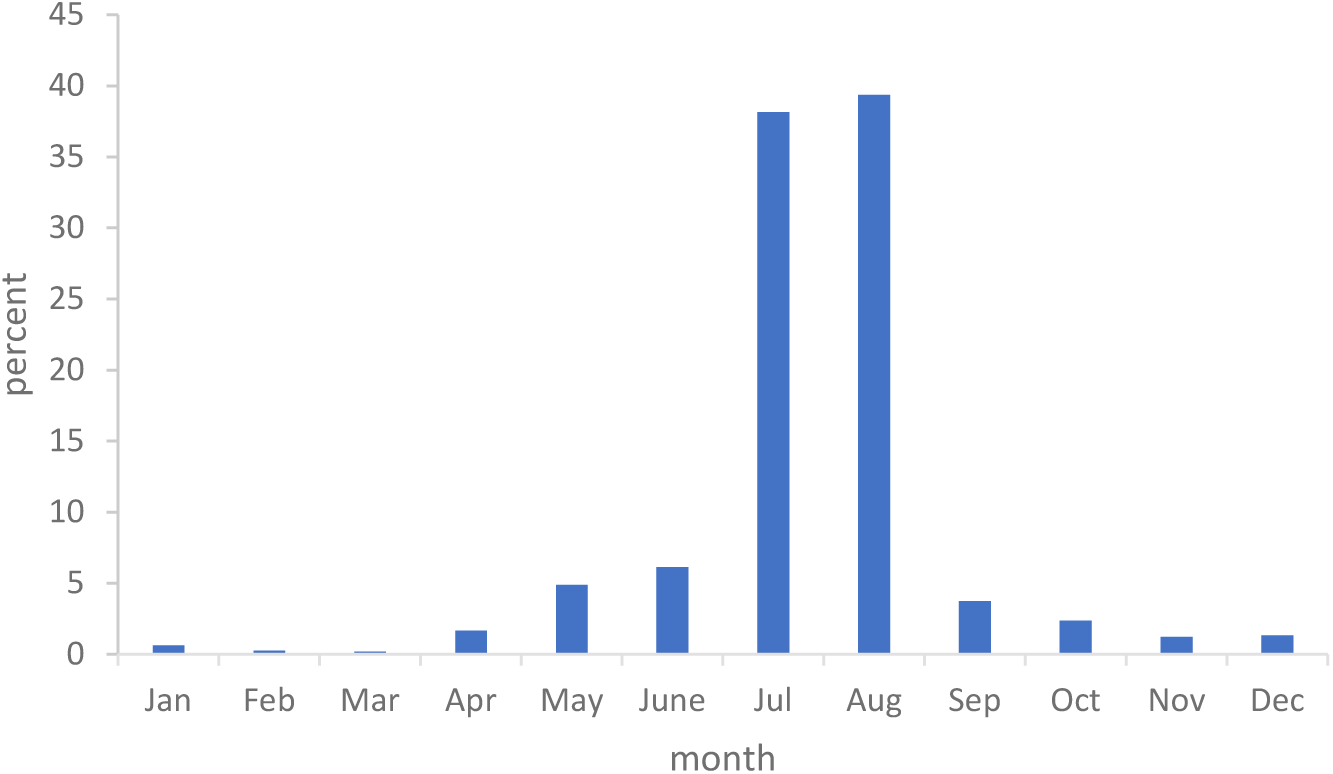
Distribution of interview months in 2010, 2014 and 2018.

**Figure A4.**
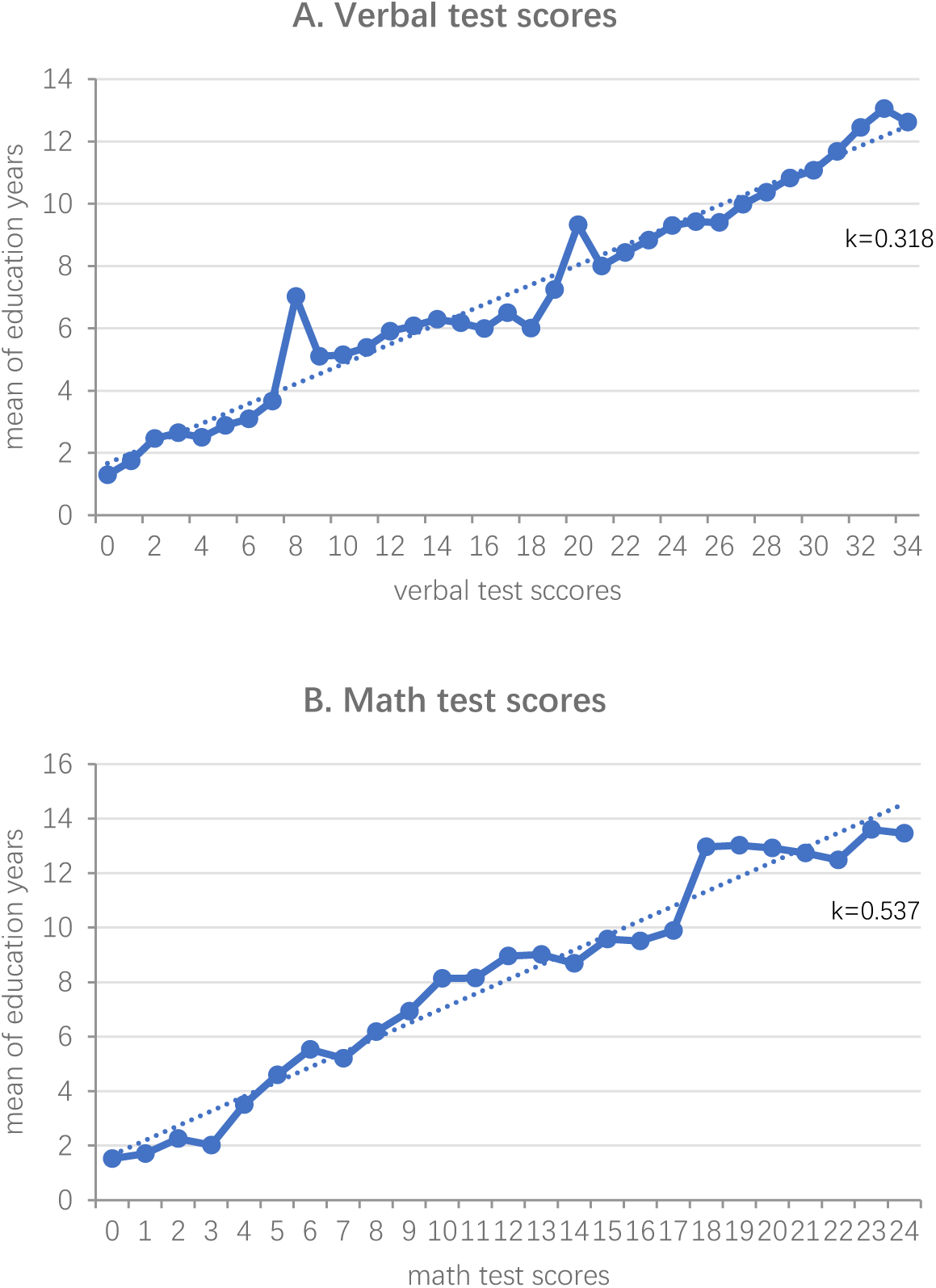
Relationship between cognitive test scores and mean values of education years. Note: k values indicate the coefficients from regressing mean values of education years on verbal test scores/math test scores.

**Figure A5.**
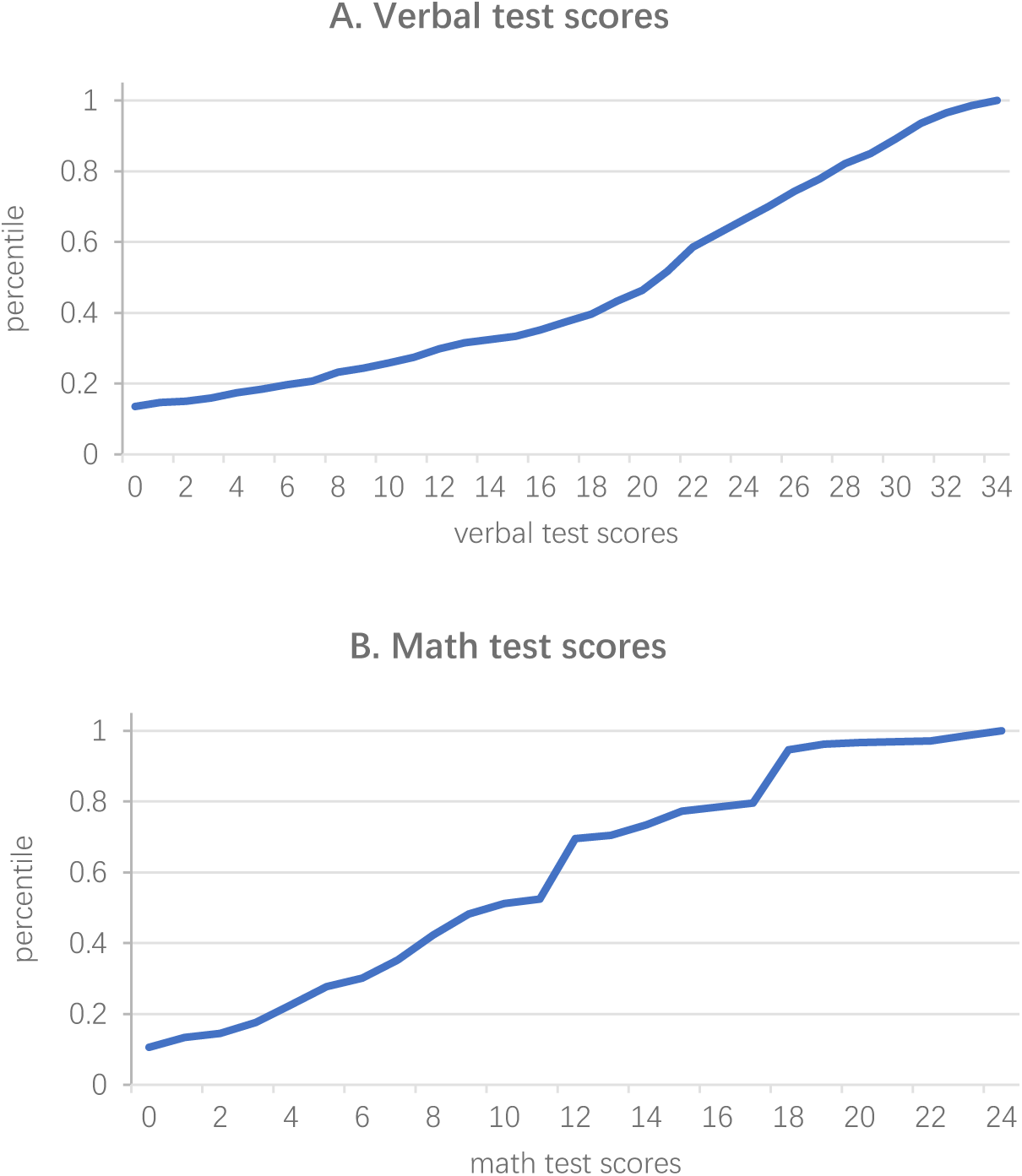
Percentiles of cognitive test scores.

**Figure A6.**
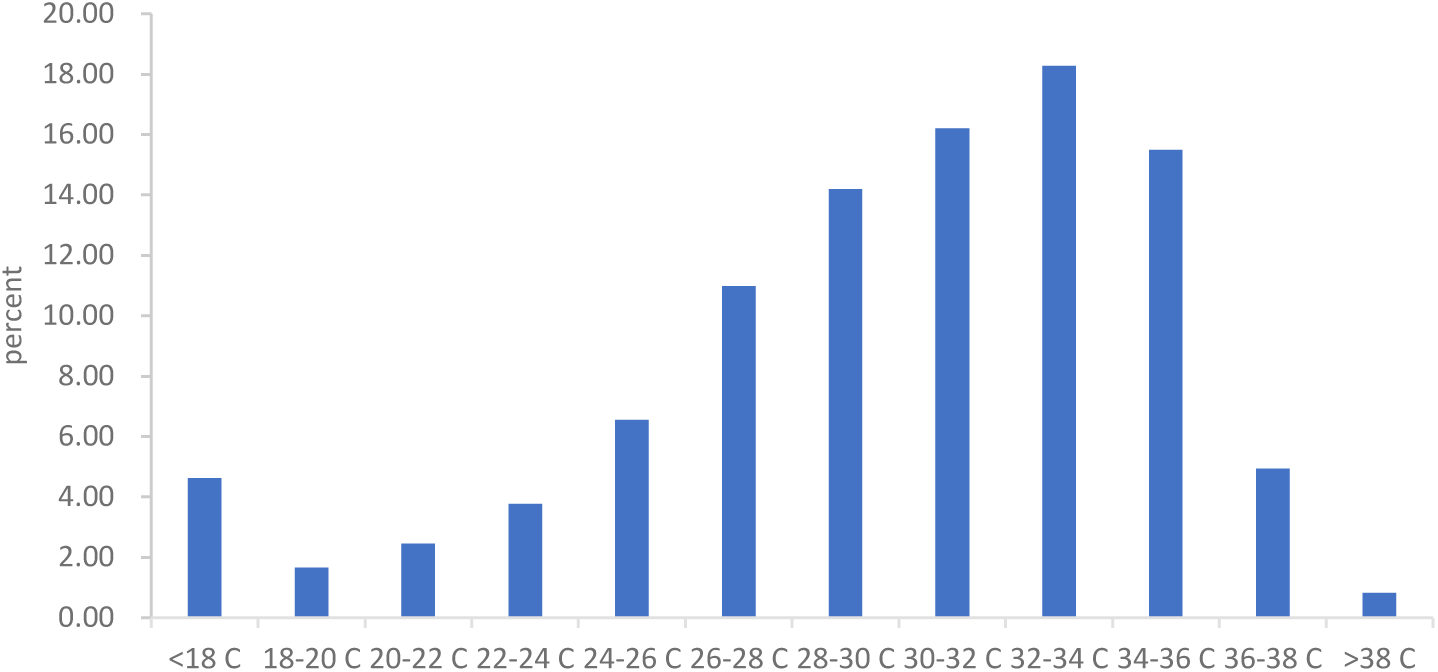
Distribution of daily maximum temperature (°C) on the test date.

**Figure A7.**
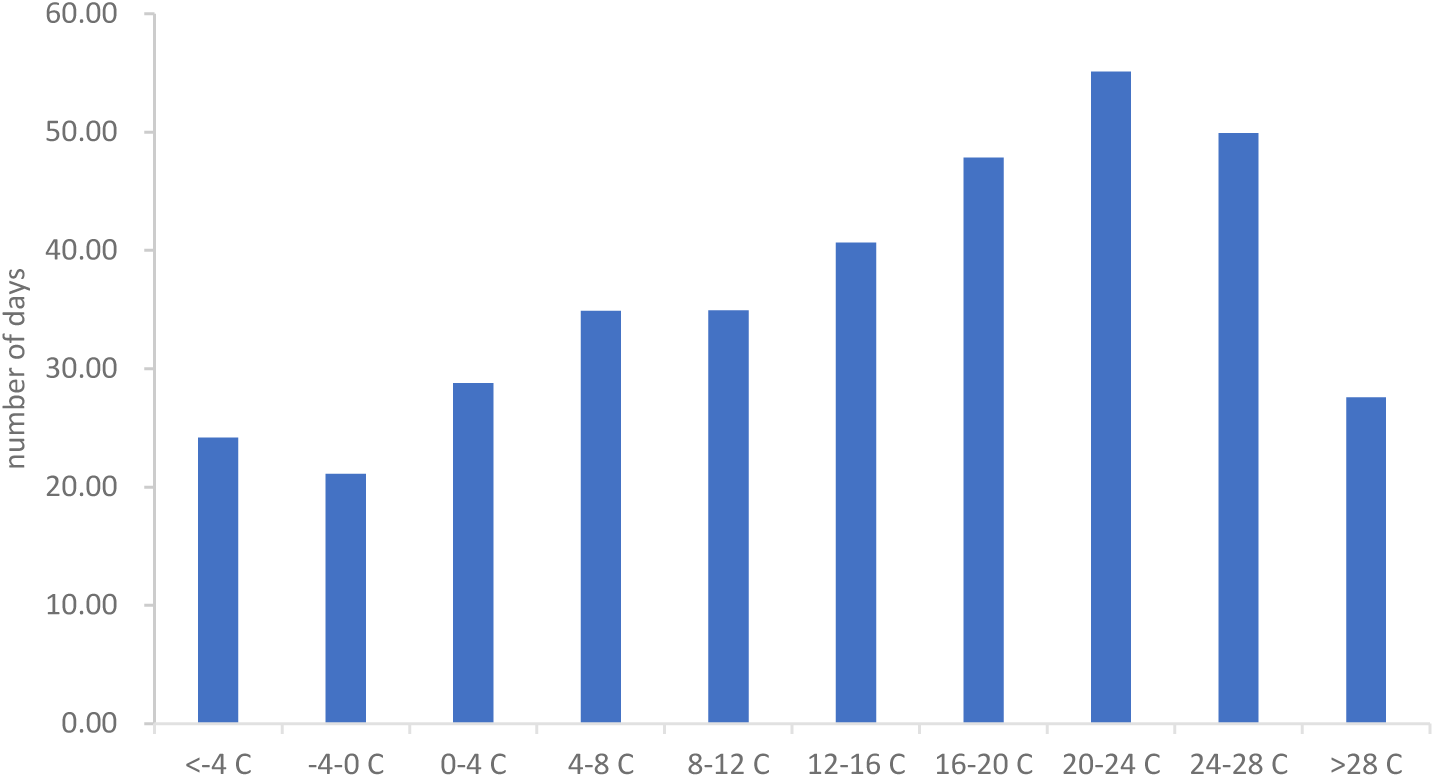
Distribution of mean temperature (°C) in the past year.

**Table A1.** Transitory effects of temperature on cognitive test scores.

| Dependent variable | A. verbal test scores |  |  | B. math test scores |  |  |
| --- | --- | --- | --- | --- | --- | --- |
|  | (1) | (2) | (3) | (4) | (5) | (6) |
| the **indicators** on the test date |  |  |  |  |  |  |
| <12 °C | -1.305***<br>(0.466) | -0.119<br>(0.481) | -0.144<br>(0.469) | 0.047<br>(0.173) | 0.065<br>(0.208) | 0.068<br>(0.224) |
| 12-14 °C | 0.197<br>(0.600) | 0.727<br>(0.444) | 0.706<br>(0.453) | 0.135<br>(0.277) | 0.246<br>(0.287) | 0.269<br>(0.298) |
| 14-16 °C | -0.126<br>(0.405) | 0.122<br>(0.424) | 0.126<br>(0.423) | -0.162<br>(0.151) | -0.109<br>(0.190) | -0.094<br>(0.194) |
| 16-18 °C | 0.081<br>(0.260) | 0.319<br>(0.255) | 0.330<br>(0.263) | -0.090<br>(0.153) | -0.064<br>(0.167) | -0.051<br>(0.168) |
| 18-20 °C | 0.042<br>(0.231) | 0.098<br>(0.214) | 0.118<br>(0.209) | -0.105<br>(0.117) | -0.027<br>(0.124) | -0.015<br>(0.126) |
| 20-22 °C | 0.104<br>(0.172) | 0.028<br>(0.172) | 0.039<br>(0.174) | -0.131<br>(0.091) | -0.084<br>(0.091) | -0.077<br>(0.094) |
| 22-24 °C |  |  |  |  |  |  |
| 24-26 °C | -0.124<br>(0.131) | -0.106<br>(0.122) | -0.104<br>(0.122) | -0.036<br>(0.079) | -0.069<br>(0.078) | -0.078<br>(0.076) |
| 26-28 °C | 0.323*<br>(0.177) | 0.262*<br>(0.155) | 0.257<br>(0.167) | -0.008<br>(0.086) | -0.062<br>(0.092) | -0.077<br>(0.095) |
| 28-30 °C | 0.307<br>(0.221) | 0.104<br>(0.183) | 0.076<br>(0.210) | -0.072<br>(0.098) | -0.112<br>(0.108) | -0.132<br>(0.124) |
| 30-32 °C | 0.497*<br>(0.294) | 0.189<br>(0.246) | 0.171<br>(0.293) | -0.044<br>(0.122) | -0.084<br>(0.127) | -0.108<br>(0.148) |
| >32 °C | 0.616<br>(0.465) | 0.003<br>(0.408) | -0.003<br>(0.444) | -0.465**<br>(0.216) | -0.499**<br>(0.225) | -0.557**<br>(0.246) |
| environmental controls | No | No | Yes | No | No | Yes |
| individual fixed effects | Yes | Yes | Yes | Yes | Yes | Yes |
| county fixed effects | No | Yes | Yes | No | Yes | Yes |
| year, month, day-of-week fixed effects | No | Yes | Yes | No | Yes | Yes |
| Observations | 70,687 | 70,687 | 70,687 | 70,687 | 70,687 | 70,687 |
| Adjusted *R*-squared | 0.770 | 0.775 | 0.775 | 0.785 | 0.785 | 0.785 |

Note: The left-out temperature bin is 22-24 °C. The coefficients can be interpreted as effects of a day in the corresponding temperature bin on cognitive test scores relative to the reference temperature category. Demographic controls include age and its square term. Environmental controls include air pollution index (API), total precipitation, wind speed, sunshine duration, and relative humidity in quadratic forms. Robust standard errors, clustered at the county level, are presented in parentheses. \* 10% significance level; \*\* 5% significance level; \*\*\* 1% significance level.

**Table A2.** Transitory effects of temperature on cognitive test scores, by AC ownership and region.

| Dependent variable | AC ownership |  |  |  | region |  |  |  |
| --- | --- | --- | --- | --- | --- | --- | --- | --- |
|  | verbal test scores |  | math test scores |  | verbal test scores |  | math test scores |  |
|  | with AC | without AC | with AC | without AC | cool | hot | cool | hot |
|  | (1) | (2) | (3) | (4) | (5) | (6) | (7) | (8) |
| temperature bins |  |  |  |  |  |  |  |  |
| <12 °C | -1.084<br>(0.777) | 0.252<br>(0.753) | -0.174<br>(0.503) | 0.502<br>(0.334) | -0.324<br>(0.658) | -1.712*<br>(0.989) | 0.255<br>(0.309) | -0.284<br>(0.482) |
| 12-14 °C | 1.037<br>(0.649) | 0.844<br>(0.685) | 0.539<br>(0.382) | 0.356<br>(0.489) | 0.285<br>(0.575) | 0.287<br>(0.797) | -0.339<br>(0.469) | 0.471<br>(0.447) |
| 14-16 °C | 0.218<br>(0.513) | 0.511<br>(0.578) | 0.011<br>(0.338) | 0.110<br>(0.246) | 0.004<br>(0.597) | -0.446<br>(0.658) | -0.283<br>(0.272) | -0.119<br>(0.261) |
| 16-18 °C | 0.242<br>(0.416) | 0.591*<br>(0.328) | 0.119<br>(0.266) | -0.023<br>(0.204) | 0.526*<br>(0.287) | -0.426<br>(0.500) | -0.143<br>(0.208) | -0.020<br>(0.271) |
| 18-20 °C | 0.621<br>(0.437) | -0.032<br>(0.267) | 0.445**<br>(0.203) | -0.128<br>(0.155) | -0.017<br>(0.236) | 0.340<br>(0.416) | -0.241<br>(0.146) | 0.455**<br>(0.210) |
| 20-22 °C | 0.254<br>(0.265) | -0.028<br>(0.216) | 0.129<br>(0.170) | -0.110<br>(0.102) | -0.026<br>(0.187) | 0.328<br>(0.316) | -0.173*<br>(0.101) | 0.224<br>(0.176) |
| 22-24 °C |  |  |  |  |  |  |  |  |
| 24-26 °C | 0.326<br>(0.234) | -0.249*<br>(0.149) | -0.004<br>(0.132) | -0.103<br>(0.088) | -0.097<br>(0.137) | -0.050<br>(0.257) | -0.108<br>(0.096) | 0.030<br>(0.121) |
| 26-28 °C | 0.695**<br>(0.313) | 0.164<br>(0.211) | 0.026<br>(0.152) | -0.083<br>(0.106) | 0.234<br>(0.180) | 0.405<br>(0.315) | -0.180*<br>(0.100) | 0.141<br>(0.181) |
| 28-30 °C | 0.463<br>(0.405) | -0.037<br>(0.221) | 0.049<br>(0.193) | -0.097<br>(0.122) | 0.199<br>(0.222) | 0.163<br>(0.383) | -0.164<br>(0.158) | 0.107<br>(0.208) |
| 30-32 °C | 0.530<br>(0.543) | -0.067<br>(0.290) | 0.073<br>(0.246) | -0.061<br>(0.144) | 0.372<br>(0.323) | 0.203<br>(0.477) | -0.114<br>(0.199) | 0.134<br>(0.244) |
| >32 °C | 0.147<br>(0.560) | 0.193<br>(0.671) | -0.364<br>(0.316) | -0.723**<br>(0.316) | 1.029<br>(1.195) | -0.248<br>(0.504) | -1.308**<br>(0.566) | -0.312<br>(0.314) |
| Observations | 20,888 | 32,641 | 20,888 | 32,641 | 33,893 | 36,794 | 33,893 | 36,794 |
| Adjusted R-squared | 0.746 | 0.787 | 0.786 | 0.788 | 0.793 | 0.762 | 0.791 | 0.780 |

Note: The left-out temperature bin is 22-24 °C. The coefficients can be interpreted as effects of a day in the corresponding temperature bin on cognitive test scores relative to the reference temperature category. All the regressions include individual fixed effects, county fixed effects, year, month, and day-of-week fixed effects. Demographic controls include age and its square term. Environmental controls include air pollution index (API), total precipitation, wind speed, sunshine duration, and relative humidity in quadratic forms. Robust standard errors, clustered at the county level, are presented in parentheses. \* 10% significance level; \*\* 5% significance level; \*\*\* 1% significance level.

**Table A3.**
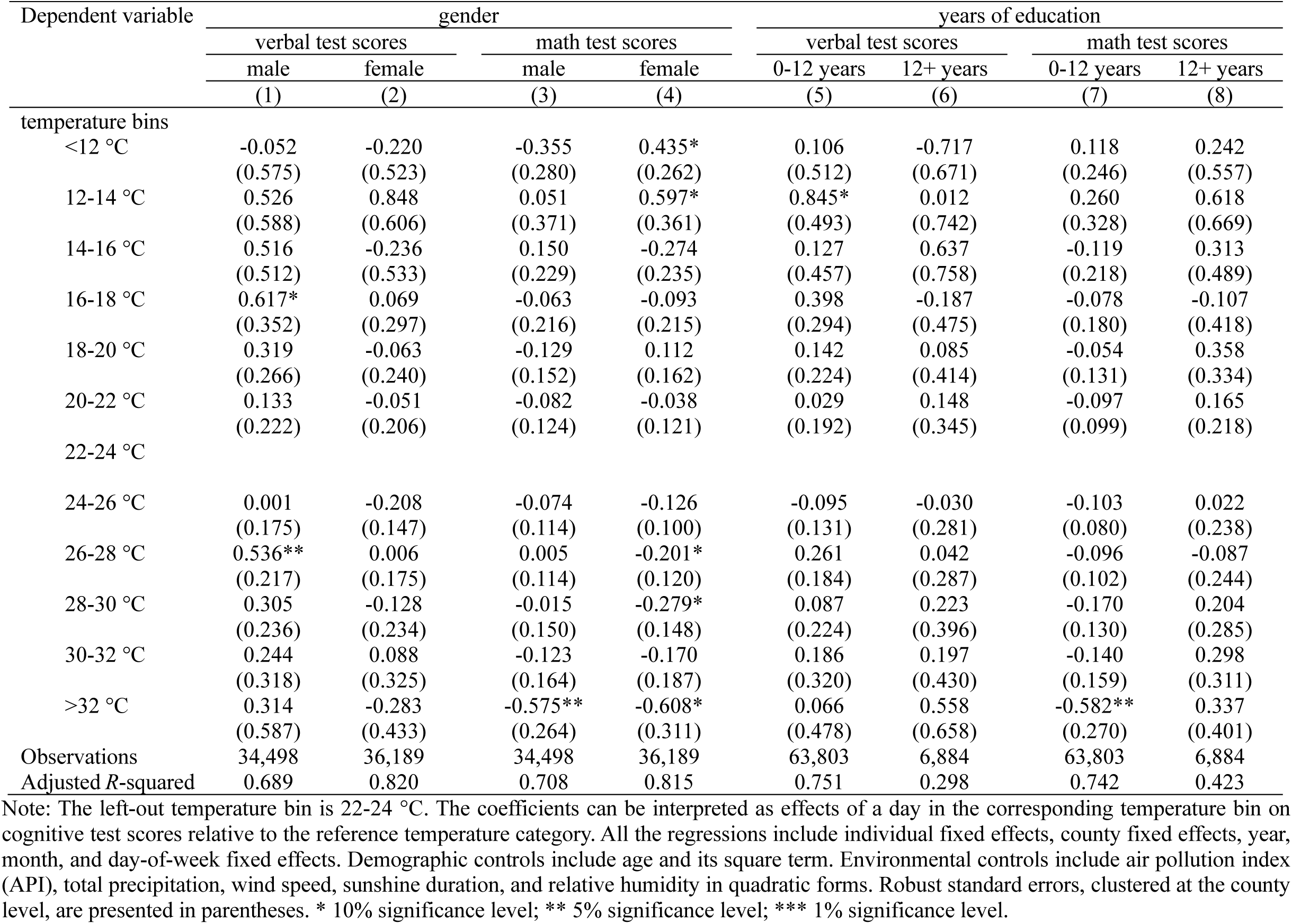
Transitory effects of temperature on cognitive test scores, by education level and gender.

| Dependent variable | gender |  |  |  | years of education |  |  |  |
| --- | --- | --- | --- | --- | --- | --- | --- | --- |
|  | verbal test scores |  | math test scores |  | verbal test scores |  | math test scores |  |
|  | male | female | male | female | 0-12 years | 12+ years | 0-12 years | 12+ years |
|  | (1) | (2) | (3) | (4) | (5) | (6) | (7) | (8) |
| temperature bins |  |  |  |  |  |  |  |  |
| <12 °C | -0.052<br>(0.575) | -0.220<br>(0.523) | -0.355<br>(0.280) | 0.435*<br>(0.262) | 0.106<br>(0.512) | -0.717<br>(0.671) | 0.118<br>(0.246) | 0.242<br>(0.557) |
| 12-14 °C | 0.526<br>(0.588) | 0.848<br>(0.606) | 0.051<br>(0.371) | 0.597*<br>(0.361) | 0.845*<br>(0.493) | 0.012<br>(0.742) | 0.260<br>(0.328) | 0.618<br>(0.669) |
| 14-16 °C | 0.516<br>(0.512) | -0.236<br>(0.533) | 0.150<br>(0.229) | -0.274<br>(0.235) | 0.127<br>(0.457) | 0.637<br>(0.758) | -0.119<br>(0.218) | 0.313<br>(0.489) |
| 16-18 °C | 0.617*<br>(0.352) | 0.069<br>(0.297) | -0.063<br>(0.216) | -0.093<br>(0.215) | 0.398<br>(0.294) | -0.187<br>(0.475) | -0.078<br>(0.180) | -0.107<br>(0.418) |
| 18-20 °C | 0.319<br>(0.266) | -0.063<br>(0.240) | -0.129<br>(0.152) | 0.112<br>(0.162) | 0.142<br>(0.224) | 0.085<br>(0.414) | -0.054<br>(0.131) | 0.358<br>(0.334) |
| 20-22 °C | 0.133<br>(0.222) | -0.051<br>(0.206) | -0.082<br>(0.124) | -0.038<br>(0.121) | 0.029<br>(0.192) | 0.148<br>(0.345) | -0.097<br>(0.099) | 0.165<br>(0.218) |
| 22-24 °C |  |  |  |  |  |  |  |  |
| 24-26 °C | 0.001<br>(0.175) | -0.208<br>(0.147) | -0.074<br>(0.114) | -0.126<br>(0.100) | -0.095<br>(0.131) | -0.030<br>(0.281) | -0.103<br>(0.080) | 0.022<br>(0.238) |
| 26-28 °C | 0.536**<br>(0.217) | 0.006<br>(0.175) | 0.005<br>(0.114) | -0.201*<br>(0.120) | 0.261<br>(0.184) | 0.042<br>(0.287) | -0.096<br>(0.102) | -0.087<br>(0.244) |
| 28-30 °C | 0.305<br>(0.236) | -0.128<br>(0.234) | -0.015<br>(0.150) | -0.279*<br>(0.148) | 0.087<br>(0.224) | 0.223<br>(0.396) | -0.170<br>(0.130) | 0.204<br>(0.285) |
| 30-32 °C | 0.244<br>(0.318) | 0.088<br>(0.325) | -0.123<br>(0.164) | -0.170<br>(0.187) | 0.186<br>(0.320) | 0.197<br>(0.430) | -0.140<br>(0.159) | 0.298<br>(0.311) |
| >32 °C | 0.314<br>(0.587) | -0.283<br>(0.433) | -0.575**<br>(0.264) | -0.608*<br>(0.311) | 0.066<br>(0.478) | 0.558<br>(0.658) | -0.582**<br>(0.270) | 0.337<br>(0.401) |
| Observations | 34,498 | 36,189 | 34,498 | 36,189 | 63,803 | 6,884 | 63,803 | 6,884 |
| Adjusted R-squared | 0.689 | 0.820 | 0.708 | 0.815 | 0.751 | 0.298 | 0.742 | 0.423 |

Note: The left-out temperature bin is 22-24 °C. The coefficients can be interpreted as effects of a day in the corresponding temperature bin on cognitive test scores relative to the reference temperature category. All the regressions include individual fixed effects, county fixed effects, year, month, and day-of-week fixed effects. Demographic controls include age and its square term. Environmental controls include air pollution index (API), total precipitation, wind speed, sunshine duration, and relative humidity in quadratic forms. Robust standard errors, clustered at the county level, are presented in parentheses. \* 10% significance level; \*\* 5% significance level; \*\*\* 1% significance level.

**Table A4.** Transitory effects of temperature on cognitive test scores, by age cohort.

| Dependent variable | verbal test scores |  |  | math test scores |  |  |
| --- | --- | --- | --- | --- | --- | --- |
|  | 0-30 | 31-59 | 60+ | 0-30 | 31-59 | 60+ |
|  | (1) | (2) | (3) | (4) | (5) | (6) |
| temperature bins |  |  |  |  |  |  |
| <12 °C | -0.811<br>(0.835) | 0.257<br>(0.502) | -0.396<br>(0.857) | 0.267<br>(0.437) | 0.002<br>(0.283) | 0.191<br>(0.388) |
| 12-14 °C | 1.841<br>(1.261) | 0.579<br>(0.594) | 0.458<br>(0.660) | 1.424**<br>(0.619) | 0.006<br>(0.392) | 0.332<br>(0.374) |
| 14-16 °C | 0.204<br>(0.918) | 0.134<br>(0.435) | 0.088<br>(0.630) | 0.146<br>(0.381) | -0.222<br>(0.222) | 0.105<br>(0.336) |
| 16-18 °C | -0.094<br>(0.587) | 0.494<br>(0.320) | 0.417<br>(0.460) | 0.002<br>(0.372) | -0.042<br>(0.179) | -0.010<br>(0.233) |
| 18-20 °C | 0.249<br>(0.394) | 0.147<br>(0.267) | 0.030<br>(0.304) | 0.210<br>(0.269) | -0.199<br>(0.161) | 0.285<br>(0.174) |
| 20-22 °C | 0.429*<br>(0.246) | -0.080<br>(0.205) | 0.138<br>(0.361) | -0.010<br>(0.197) | -0.064<br>(0.137) | -0.019<br>(0.168) |
| 22-24 °C |  |  |  |  |  |  |
| 24-26 °C | 0.165<br>(0.222) | -0.217<br>(0.166) | 0.006<br>(0.220) | -0.030<br>(0.175) | -0.100<br>(0.091) | 0.118<br>(0.130) |
| 26-28 °C | 0.635**<br>(0.265) | -0.029<br>(0.198) | 0.594**<br>(0.269) | 0.035<br>(0.177) | -0.212*<br>(0.115) | 0.144<br>(0.164) |
| 28-30 °C | 0.336<br>(0.377) | 0.147<br>(0.264) | -0.092<br>(0.299) | 0.288<br>(0.233) | -0.282*<br>(0.145) | 0.091<br>(0.184) |
| 30-32 °C | 0.734<br>(0.508) | 0.135<br>(0.330) | -0.041<br>(0.365) | 0.144<br>(0.251) | -0.202<br>(0.173) | 0.031<br>(0.219) |
| >32 °C | 0.532<br>(0.752) | 0.100<br>(0.532) | -0.335<br>(0.478) | 0.544<br>(0.431) | -0.564*<br>(0.292) | -0.778**<br>(0.331) |
| Observations | 18,615 | 35,688 | 16,384 | 18,615 | 35,688 | 16,384 |
| Adjusted R-squared | 0.550 | 0.756 | 0.748 | 0.694 | 0.784 | 0.758 |

Note: The left-out temperature bin is 22-24 °C. The coefficients can be interpreted as effects of a day in the corresponding temperature bin on cognitive test scores relative to the reference temperature category. All the regressions include individual fixed effects, county fixed effects, year, month, and day-of-week fixed effects. Demographic controls include age and its square term. Environmental controls include air pollution index (API), total precipitation, wind speed, sunshine duration, and relative humidity in quadratic forms. Robust standard errors, clustered at the county level, are presented in parentheses. \* 10% significance level; \*\* 5% significance level; \*\*\* 1% significance level.

## Footnotes

1 The survey uses multistage probability proportional to size sampling with implicit stratification to better represent Chinese society. The 2010 CFPS baseline sample is drawn through three stages (i.e., county, village, and household) from 25 provinces. The 162 randomly chosen counties largely represent Chinese society (Xie and Hu 2014).

2 Specifically, those whose education level is primary school or below start with the 1st question; those who attended middle school begin with the 9th question in the verbal test and the 13th question in the math test; and those who finished high school or above start with the 21st question in the verbal test and the 19th question in the math test.

3 Our matching radius was comparable to those used in similar studies (Deschênes and Greenstone 2007, 2011).

4 We could not plot the locations of CFPS counties in Figure A1 due to restricted data.

5 Carbon monoxide (CO), ozone, and particulate matter smaller than 2.5 micrometers (PM2.5) were not added to the basket of the index until 2013. Because all the cognitive tests were administered between 2010 and 2018, we transform the air quality index (AQI) to the API in 2014 and 2018, and use the API based on SO_2_, NO_2_, and PM10 in our paper.

6 The county fixed effects cannot be wiped out by individual fixed effects since some respondents do not live in the same counties across the three waves.

## Notes

### Competing Interest Statement

The authors have declared no competing interest.

